# Neural and psychophysical predictors of treatment response to transcranial direct current stimulation and mindfulness-based meditation for knee osteoarthritis pain

**DOI:** 10.64898/2026.02.02.26345407

**Authors:** Chiyoung Lee, Juyoung Park, Hongyu Miao, Hyochol Ahn

## Abstract

**Aim:** We investigated the heterogeneity of treatment effects in transcranial direct current stimulation (tDCS) with mindfulness-based meditation (MBM) and within each individual study group (tDCS alone, MBM alone, and sham) among individuals with symptomatic knee osteoarthritis. We also explored participant characteristics underlying this heterogeneity.

**Methods:** This secondary analysis drew on a double-blind, randomized, sham-controlled, phase II, parallel-group trial in which 200 participants were assigned to one of four groups: (1) active tDCS + active MBM, (2) active tDCS + sham MBM, (3) sham tDCS + active MBM, or (4) sham tDCS + sham MBM. Participants received ten 20-minute tDCS sessions (active or sham) administered concurrently with MBM (active or sham). Latent class growth analysis was used to identify subgroups with distinct treatment response trajectories (responders vs. non-responders) based on changes in clinical pain (Numeric Rating Scale) from baseline to post-intervention. Generalized linear models were then applied to determine baseline factors associated with participants’ response classification, including demographic, clinical, and psychological characteristics; quantitative sensory testing battery; and pain-related cortical hemodynamic activity measured using functional near-infrared spectroscopy (fNIRS) in response to punctate and thermal stimuli.

**Results:** Responders in the active tDCS + active MBM and active tDCS + sham MBM groups demonstrated greater improvements in clinical pain from baseline to post-intervention than non-responders (*p* < 0.001). In the active tDCS + active MBM group, greater cortical activation in the fNIRS channel S06-D06 of the left somatosensory cortex in response to punctate stimuli, identifying as white, and lower conditioned pain modulation (reflecting less efficient endogenous pain modulation), were significantly associated with being responders (*p* < 0.05). In the active tDCS + sham MBM group, younger age and lower heat pain tolerance at the knee were significantly associated with being responders (*p* < 0.05). No clear response patterns were observed in the remaining groups.

**Conclusion:** Factors underlying heterogeneity of treatment effects, including somatosensory cortical activation and pain modulatory profiles, may provide preliminary insights to inform the development of personalized neuromodulation (stimulation) protocols.

## 1. Introduction

As evidence increasingly supports centrally mediated mechanisms underlying chronic pain in knee osteoarthritis (KOA), interest has grown in approaches that target pain-related brain function, especially transcranial direct current stimulation (tDCS). tDCS improves pain system function by modulating a wide neural network involved in pain processing, via transcranial electrical field stimulation, including the thalamic nuclei, limbic system, brainstem nuclei, and spinal cord.^1–6^ The efficacy of tDCS in alleviating KOA pain and symptoms has been demonstrated in several clinical trials.^7–11^ Likewise, mindfulness-based meditation (MBM) has been shown to improve pain-related brain function^12–16^ and relieve pain in multiple chronic pain conditions.^17–19^ Since tDCS promotes neuroplasticity,^20,21^ this neuromodulatory effect may potentiate the effects of MBM, which also encourages adaptive changes in the brain, thereby further reducing clinical pain.^22^ Consequently, a combined approach of tDCS and MBM has emerged as a multimodal strategy for improving clinical outcomes in KOA.^22^

While the overall summary result from randomized trials, known as the average treatment effect, has long guided evidence-based clinical decisions, there is growing recognition that treatment responses vary across individuals. This concept is described as heterogeneity of treatment effects (HTE)^23^ and refers to non-random variation in the magnitude or direction of the treatment effect on a clinical outcome of interest across patient subgroups defined by one or more covariates.^24^ Recent studies have demonstrated HTE in response to tDCS by identifying high- and low-responder groups and have reported characteristics underlying this variability, including educational attainment, body mass index (BMI), pain sensitivity, and psychological factors.^25–27^ Similarly, individual variation in response to MBM may exist, with some participants exhibiting greater or reduced meaningful improvement. Indeed, understanding the characteristics that make individuals more or less likely to benefit from treatment is essential for improved insights into patient selection and interpretation in future clinical trials, ensuring optimal treatment administration, and ultimately helping to develop personalized treatment plans.

To our knowledge, no studies have examined the HTE in individuals with KOA undergoing combined tDCS and MBM treatment. This study represents a secondary analysis of a double-blind, randomized, sham-controlled phase II trial with a 2 × 2 factorial design (1:1:1:1 allocation) that evaluated remotely supervised, home-based tDCS combined with MBM in 200 older adults with symptomatic KOA. Our primary aim was to assess HTE for the main intervention (tDCS + MBM) and identify baseline characteristics underlying this heterogeneity. Additionally, we conducted the same analyses for the three remaining treatment arms (tDCS only, MBM only, and sham) to achieve a more comprehensive understanding of response patterns across modalities. Specifically, this study differed from prior investigations of HTE in neuromodulation ^25–27^ by incorporating an expanded set of baseline variables, including demographic, clinical, and psychological characteristics; a multimodal quantitative sensory testing (QST) battery; neuroimaging-derived measures obtained from functional near-infrared spectroscopy (fNIRS) to quantify neural markers of pain processing at the central nervous system level underlying HTE.

## 2. Methods

### 2.1. Study Design

This is a secondary analysis of data from a double-blind, randomized, sham-controlled, phase II, 2 × 2 factorial, parallel-group trial. This study involved a two-week, home-based tDCS and MBM intervention, monitored in real time via secure videoconferencing. A total of 200 participants completed the study following attrition (*n* = 8), all of whom provided written informed consent and were randomly assigned to one of four groups: (1) active tDCS + active MBM, (2) active tDCS + sham MBM, (3) sham tDCS + active MBM, or (4) sham tDCS + sham MBM (**Figure 1**). Randomization was conducted based on the order of participant enrollment using a pre-generated list created in SAS© software, Version 9.4 (SAS Institute Inc., Cary, NC) by a statistician who was not involved in the trial’s clinical components. To ensure a balanced distribution across groups, covariate-adaptive randomization was implemented.

**Figure 1.**
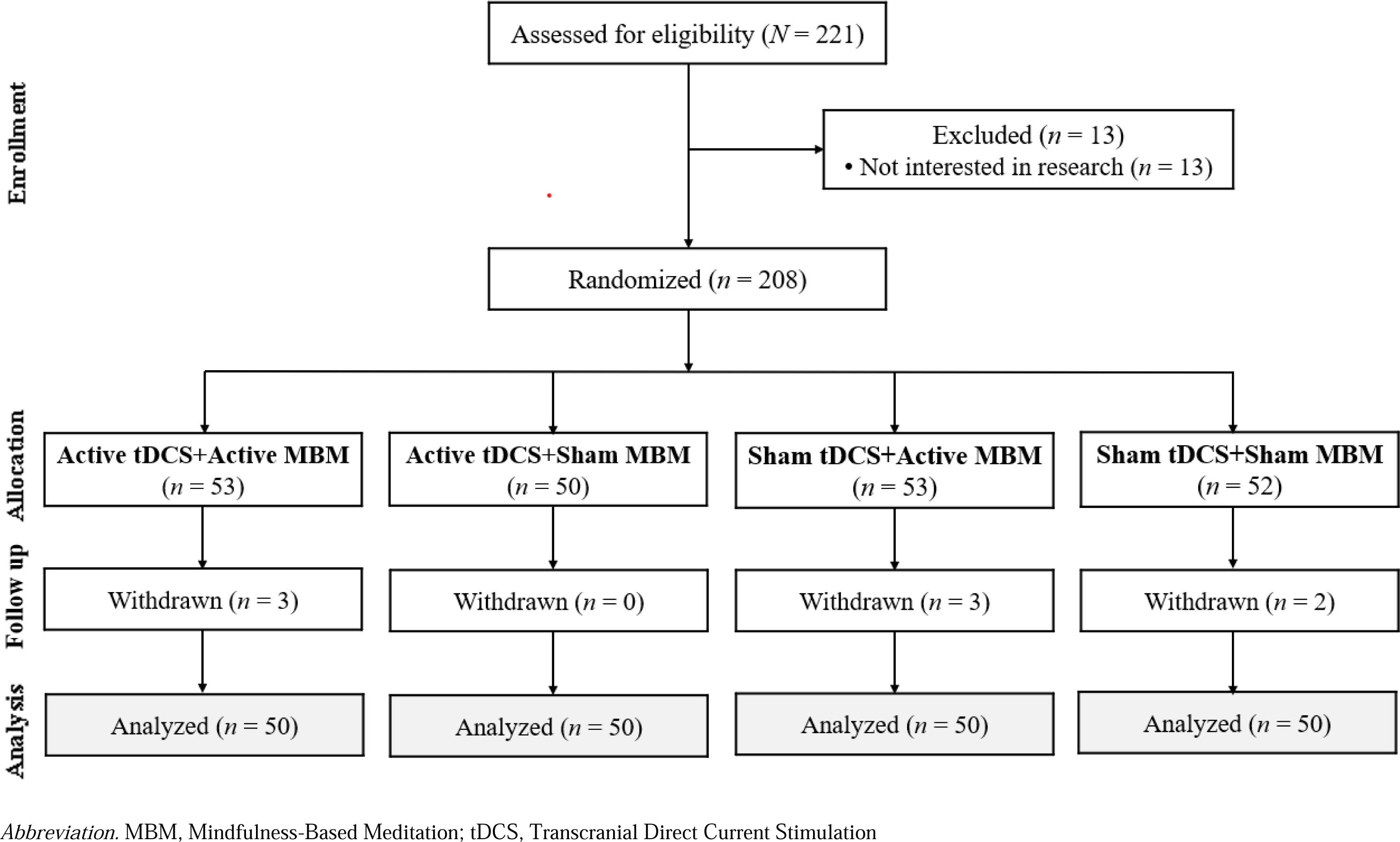
Clinical trial flow diagram.

The trial was approved by the University of Arizona Institutional Review Board (STUDY00003164) and registered at ClinicalTrials.gov (Identifier: NCT04375072; First posted: May 5, 2020). Written informed consent was obtained from all participants in accordance with the Declaration of Helsinki. The full trial protocol and statistical analysis plan can be accessed through the ClinicalTrials.gov study record (https://clinicaltrials.gov/study/NCT04375072). Recruitment occurred from August 2020 to January 2025. Participants completed follow-up assessments through 3 months post-intervention.

### 2.2. Participants

Eligible participants included adults aged 50–85 years who satisfied the American College of Rheumatology (ACR) clinical criteria for symptomatic KOA.^28^ Additional inclusion criteria included the following: (1) participants experienced KOA pain within the past 3 months with an average numeric rating scale (NRS) score of ≥ 30 on a 0–100 scale; (2) participants demonstrated adequate proficiency in English; (3) no expected changes in pain medications during the study period. The ACR clinical criteria stipulated that at least three of the following were present: (a) age > 50 years, (b) morning stiffness lasting < 30 minutes, (c) crepitus, (d) bony tenderness, (e) bony enlargement, or (f) absence of palpable warmth. The key exclusion criteria included the following: (1) a history of brain surgery, tumors, seizures, stroke, epilepsy, or intracranial metal implants; (2) systemic rheumatologic disease (e.g., rheumatoid arthritis); (3) cognitive impairment (MMSE ≤ 23); (4) prior prosthetic knee replacement or non-arthroscopic knee surgery; or (5) insufficient access to a device capable of secure video conferencing.

### 2.3. Intervention

Participants received ten sessions of anodal tDCS (active or sham) simultaneously with MBM (active or sham). Sessions were conducted and monitored in real time by trained research staff via secure videoconferencing platforms, including WebEx and Zoom. At each participant interaction, staff confirmed that participants safely operate the device, tolerate the session without adverse effects, and adhere to the study schedule.

#### 2.3.1. tDCS

Active stimulation was delivered at a constant current of 2 mA for 20 minutes per session, administered daily over two weeks (Monday through Friday; 10 sessions total), using the Soterix 1×1 tDCS mini-CT stimulator (Soterix Medical Inc., NY). Headgear was used with 5 x 7 cm saline-soaked sponge electrodes to enhance reproducibility and participant comfort. The anode was placed over the M1 region (positions C3 or C4 according to the international 10/20 EEG system) contralateral to the index knee, and the cathode was positioned over the supraorbital area (Fp1 or Fp2) ipsilateral to the index knee, as per evidence-based guidelines and prior tDCS research.^7,29,30^ Participants self-administered each stimulation session after receiving a single-use unlock code from the research staff. For sham stimulation, the electrode setup was identical to the active tDCS setup, but the current was discontinued after 30 seconds.^7,31^ This approach reproduces the initial sensations of active tDCS, such as scalp itching and tingling, thereby maintaining blinding for both participants and researchers.

#### 2.3.2. MBM

MBM was administered concurrently with tDCS for 20 minutes per session, daily for two weeks, using a prerecorded audio meditation delivered via a CD. The instructions were developed and recorded by an experienced mind–body intervention specialist with over 20 years of experience. Participants followed the guided audio, which guided progressively deeper mindfulness exercises, including controlled breathing, body awareness, and the cultivation of compassion. The sham recording instructed participants to relax and take deep breaths every 3 minutes, without the specific mindfulness-based instructions, such as practicing mindful attention to the breath in a non-evaluative manner. All other aspects of the sham MBM intervention (e.g., body position, time spent listening to instructions, eyes closed) aligned with the active MBM.^32^

### 2.4. Measures

#### 2.4.1. Baseline Characteristics

Demographic characteristics, including age (continuous), BMI (kg/m²), gender (male *vs.* female), race (white *vs*. non-white), education (high school or less *vs.* college or more), and marital status (married/partnered *vs.* non-married/unpartnered), as well as clinical characteristics, such as the average duration of osteoarthritis (months) and Kellgren-Lawrence score were collected. Psychological characteristics included pain catastrophizing, depressive symptoms, and COVID stress. Pain catastrophizing was assessed using the pain catastrophizing scale (PCS), a 13-item measure comprising a set of distorted and exaggerated emotional and cognitive reactions toward pain experiences leading to increased pain-related distress and disability.^33,34^ Items were rated on a 0 (“not at all”) to 4 (“all the time”) scale, and total scores were calculated by summing all items. The Center for Epidemiological Studies-Depression (CES-D) scale was used to assess depressive symptoms over the past week.^35^ The 20-item CES-D scale rates each item from 0 to 3, yielding total scores ranging from 0 (lowest) to 60 (highest). COVID stress was measured using the COVID stress scales (CSS), a 36-item instrument with a total score ranging from 0 to 144, with higher scores indicating greater stress.^36^

The QST methods involve the administration of various stimulus modalities (e.g., thermal and mechanical) and the evaluation of various perceptual endpoints (e.g., threshold, tolerance, and suprathreshold scaling).^37^ Furthermore, approaches for assessing pain modulatory function, including both inhibition and facilitation, are increasingly used.^37^ In this study, the tests included heat pain threshold (HPTh), heat pain tolerance (HPTo), pressure pain threshold (PPTh), punctate mechanical pain, temporal summation of pain (TSP), conditioned pain modulation (CPM), and cold pain intensity. More details on QST procedures are provided in **Supplementary Material S1.**

#### 2.4.2. Treatment Outcome: Clinical Pain

The treatment outcome in this study—the numeric rating scale (NRS)—was measured at three time points: baseline, day 5, and day 10 (post-intervention). The NRS represents a valid, reliable, and responsive measure of pain widely used in individuals with KOA.^38^ Participants were asked to rate their pain on a scale of 0 to 10, with 0 indicating “no pain” and 10 indicating “the worst pain imaginable”.

#### 2.4.3. fNIRS

fNIRS is a noninvasive and cost-effective optical neuroimaging method that measures cerebral hemodynamic activity at the cortical level by passing harmless wavelengths of light through the scalp, cerebrospinal fluid, and cortical layers and detecting the light back-scattered by such tissues. fNIRS uses light at multiple wavelengths to measure changes in concentrations of oxygenated hemoglobin (HbO) and deoxygenated hemoglobin (HbR), thus providing functionally relevant insight into cortical hemodynamics and oxidative metabolism. Although fNIRS can penetrate tissues no more than 3 cm underneath the scalp, it has excellent temporal resolution (>10 Hz) and sufficient spatial resolution at the cortical level (in the order of cm^2^) to assess cortical hemodynamic activity through topographic imaging.^39^

We measured cortical responses using a continuous-wave, multichannel fNIRS imaging system (LIGHTNIRS, Shimadzu, Kyoto, Japan), which included 8 light sources (emitting at 780, 805, and 830 nm) and 8 detectors integrated into a flexible headgear. Optodes were arranged to bilaterally cover the prefrontal and somatosensory cortices, consistent with prior pain-related fNIRS studies.^40–42^ This configuration yielded 20 measurement channels, with a sampling rate of 13.3 Hz and a mean source–detector separation of 34.3 mm (standard deviation [SD] = 8 mm) (**Figure 2**).

**Figure 2.**
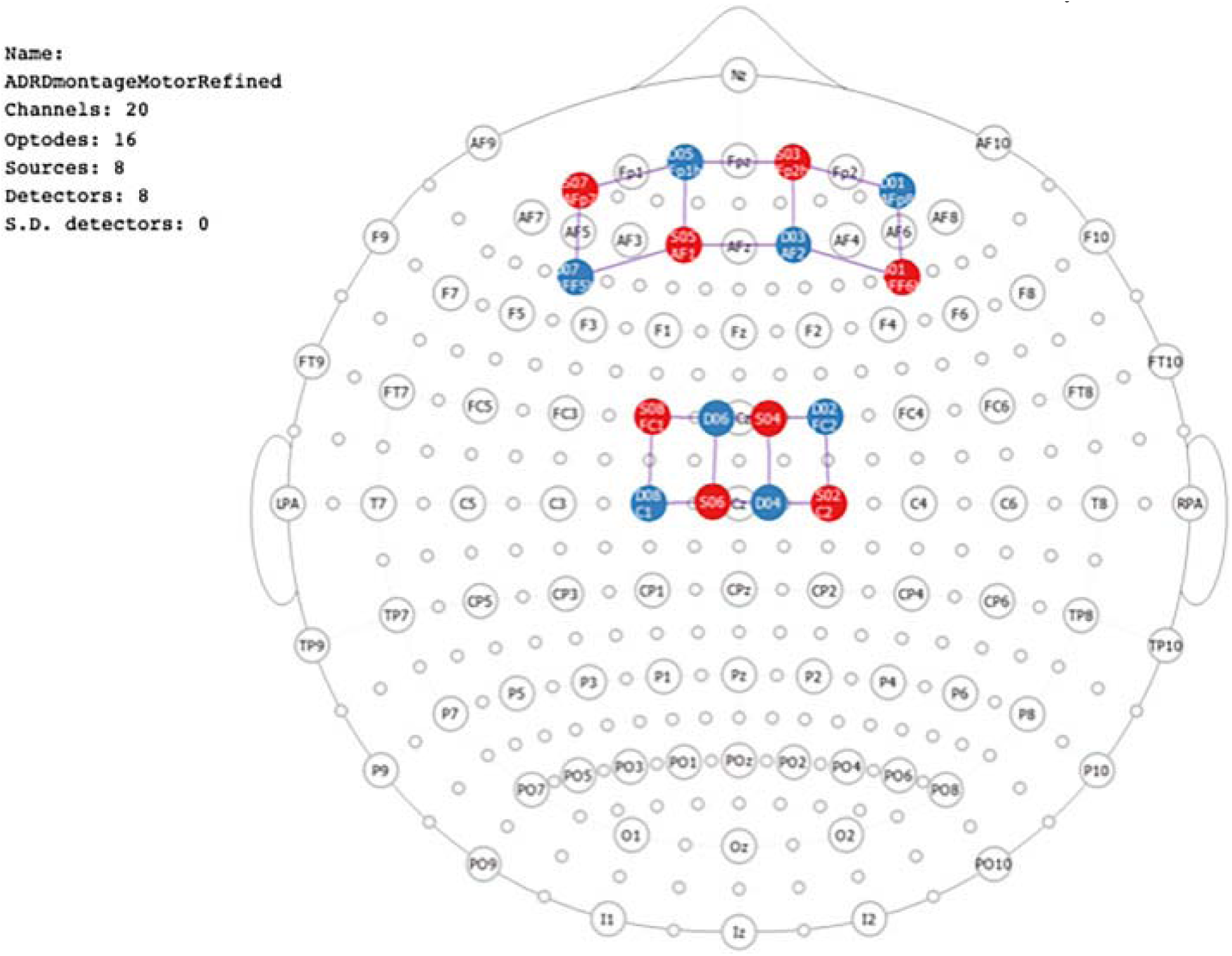
fNIRS probe design, with eight light sources (red circles) and eight light detectors (blue circles) forming 20 measurement channels (purple lines) represented over the 10–10 international system

**Figure 3.**
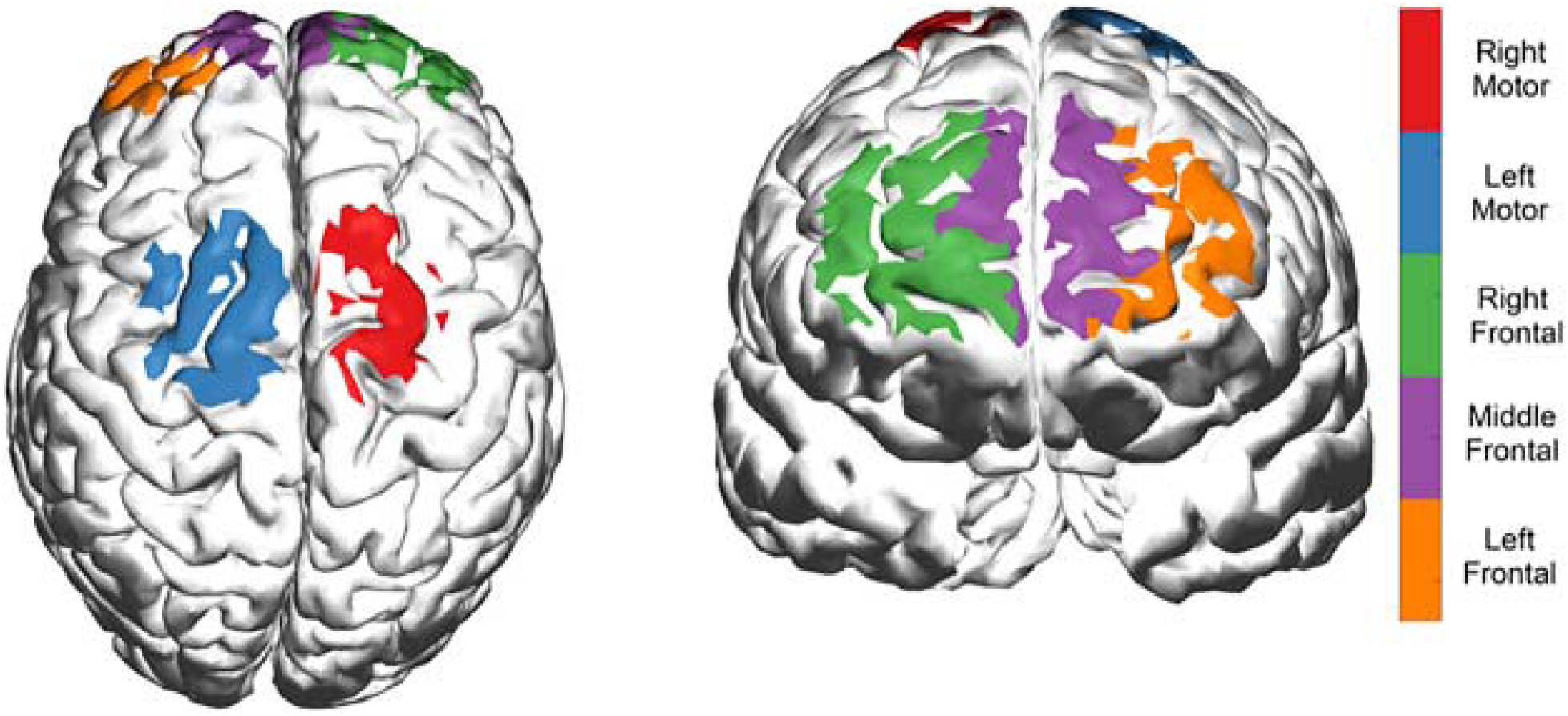
Five regions of interest covering left, middle, and right frontal and left and right motor areas.

Optical recordings were collected during pressure and thermal pain stimulation. Pressure pain was induced by periodic tapping of the symptomatic knee using a Touch Test Sensory Evaluator (North Coast Medical & Rehabilitation Products), which allowed standardized force application (X g). Thermal pain stimulation was delivered using a temperature-controlled system (Medoc TSA-II Neurosensory Analyzer) via a 16 mm ×16 mm thermode applied to the forearm. Both tasks followed a simple block design consisting of 60-second baseline rest, six cycles of 20-second stimulation with 30-second rest periods, and a final 60-second rest. Preprocessing, quality control procedures, region-of-interest definitions, and statistical modeling of fNIRS data are described in **Section 2.5.1**.

### 2.5. Statistical Analysis

Summary statistics (mean and SD) were calculated to describe participant characteristics. All analyses were conducted separately for the four study groups. To classify participants as responders or non-responders, we calculated changes in treatment outcomes (i.e., NRS) across the three time points (baseline, day 5, and day 10) and applied seven distinct methods: either (a) setting zero, mean, and median as the grouping criteria for the difference in values between baseline and the other two time points, or (b) using latent class growth analysis (LCGA), two growth mixture models (GMM1: GMM with class-specific random intercepts; GMM2: GMM with class-specific random intercepts and random slopes)^43^ and group-based multivariate trajectory modeling (GBMTM)^44^ for the differences in values between the baseline and two remaining time points. Treatment outcomes had complete follow-up data, with no missing values at any time point. Change scores between responders and non-responders were compared using the Wilcoxon rank-sum test (when normality was violated) or the *t*-test.

Associations between response groups (responders vs. non-responders) and (1) demographic, (2) clinical, (3) psychological characteristics, (4) QST measures, and (5) β coefficients from fNIRS channels/ROIs were evaluated using the Chi-square test for categorical variables and the Kruskal–Wallis test for continuous variables. Generalized linear models with backward stepwise selection based on the Bayesian information criterion (BIC) were used to identify factors significantly associated with response-group classification. Unless otherwise specified, the significance level for all tests was set to 0.05.

#### 2.5.1 Pre-processing and Analysis of fNIRS Data

Raw optical data were first evaluated for quality using a combined approach that integrated automated detection of low-quality signals with visual inspection. Specifically, optical signals from each channel were segmented into 5-second windows, bandpass filtered (0.5–2.0 Hz), and normalized at both wavelengths to isolate cardiac oscillations. Two metrics were then computed: (a) the scalp contact index (SCI), defined as the zero-lag cross-correlation between the signals and expressed as a score from 0 to 1 (with 1 indicating optimal contact), and (b) the peak power of this cross-correlation. Strong optode–scalp coupling produced raw signals with pronounced cardiac pulsations, resulting in high SCI and peak power values. Each 5-second window was classified as high quality if SCI exceeded 0.7 and peak spectral power (PSP) exceeded 0.1. Optical channels with excessive noise, defined as having less than 60% of high-quality data, were removed from further analysis. After quality assessment, raw optical data were converted to optical density. To further ensure data integrity, all remaining motion artifacts were detected and corrected using the Temporal Derivative Distribution Repair algorithm (TDDR).^45^

Processed optical density signals were then converted to HbO and HbR concentration changes over time using the modified Beer–Lambert law. The association between cortical hemodynamic activity (i.e., changes in HbO and HbR over time) and blocked thermal/punctate stimulation was quantified at the subject level (i.e., each optical channel of each subject) using a general linear model (GLM) fitted with autoregressive iteratively reweighted least squares (AR-IWLS). The GLM estimated coefficient beta (β) is a measure of hemodynamic response function in response to the pain stimuli. For ROI analysis, the resulting β values were averaged across the channels within certain ROI. The five brain ROIs are shown in **Figure 2** and were defined as 1) right dorsolateral prefrontal cortex (rdlPFC) with channels S01-D01, S01-D03, and S03-D01, 2) medial prefrontal cortex (mPFC) with channels S03-D03, S03-D05, S05-D03, and S05-D05, 3) left dorsolateral prefrontal cortex (ldlPFC) with channels S07-D05, S07-D07, and S05-D07, 4) right somatosensory cortex (rS1) with channels S02-D02, S02-D04, S04-D02, and S04-D04, and 5) left somatosensory cortex (lS1) with channels S06-D06, S06-D08, S08-D06, and S08-D08. All quality assessment algorithms were developed by our team, while data conversion and statistical analyses were performed using the AnalyzIR toolbox. All code was executed in MATLAB® (Natick, MA). For fNIRS analyses, we focused on HbO, which exhibits higher signal-to-noise ratio and greater sensitivity to task-evoked cortical activation than HbR.

## 3. Results

**Supplementary Table 1** presents the baseline characteristics of participants by study group. The average change in NRS score (ΔNRS) from baseline to day 10 was significantly lower in the sham tDCS + active MBM group (–7.16 [SD 22.24]) and the sham tDCS + sham MBM group (–6.20 [SD 17.99]) compared with the active tDCS + active MBM group (19.41 [SD 19.76]) and the active tDCS + sham MBM group (–16.78 [SD 17.90]). **Table 1** and **Figures 4–7** present the group-classification results for the first four methods (zero-, mean-, and median-change-score criteria, and LCGA). The overlap in classification across these methods is summarized in **Table 2**. The results from the GMM1, GMM2, and GBMTM approaches are not included because all three methods demonstrated inaccuracies in classifying both relatively stable trajectories and those with more pronounced trends, particularly GBMTM, which showed no clear trend in its classifications (**Supplementary Figure 1**).

**Figure 4.**
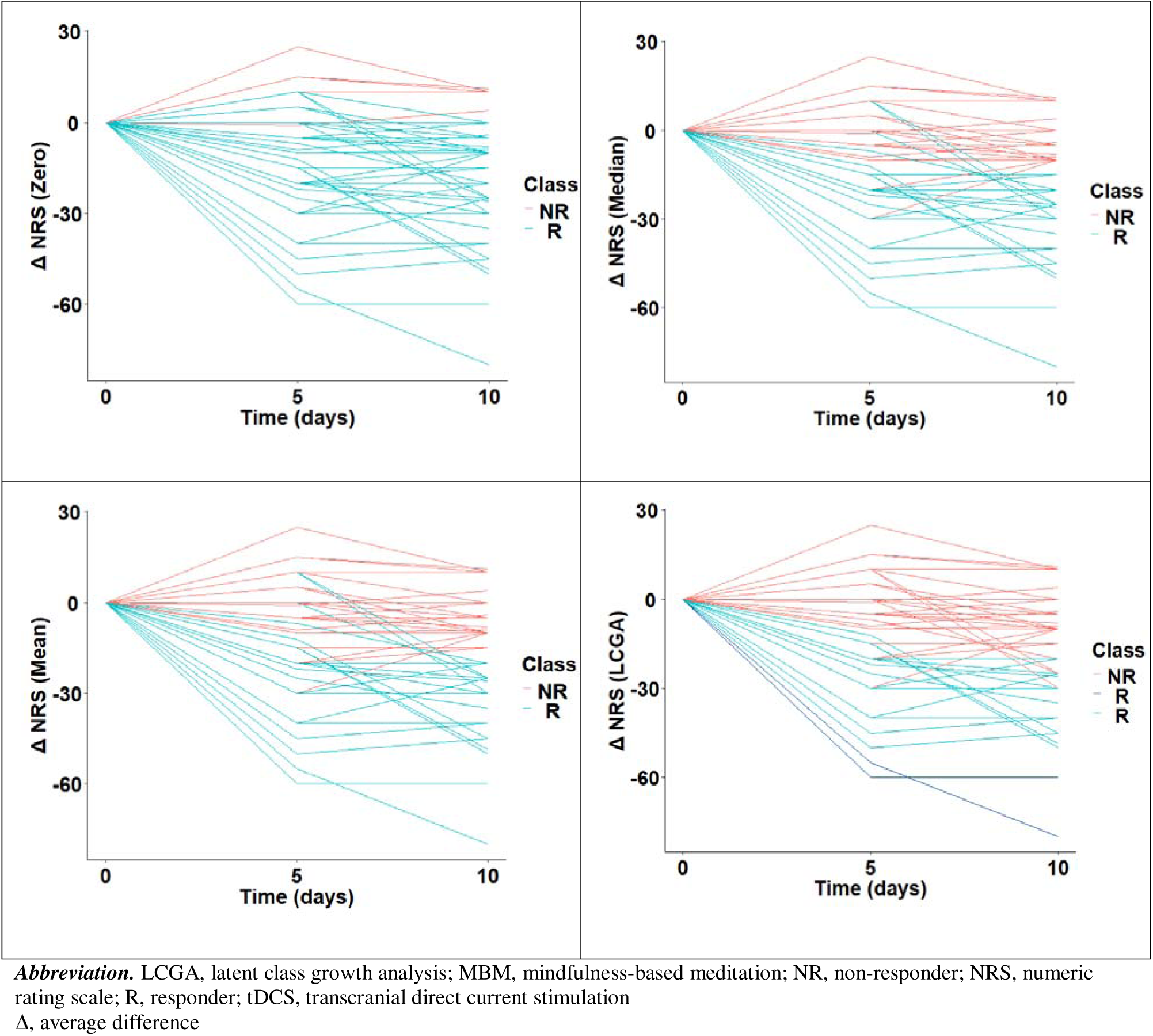
Group classification results for the first four methods: tDCS + MBM: active tDCS + active MBM.

**Figure 5.**
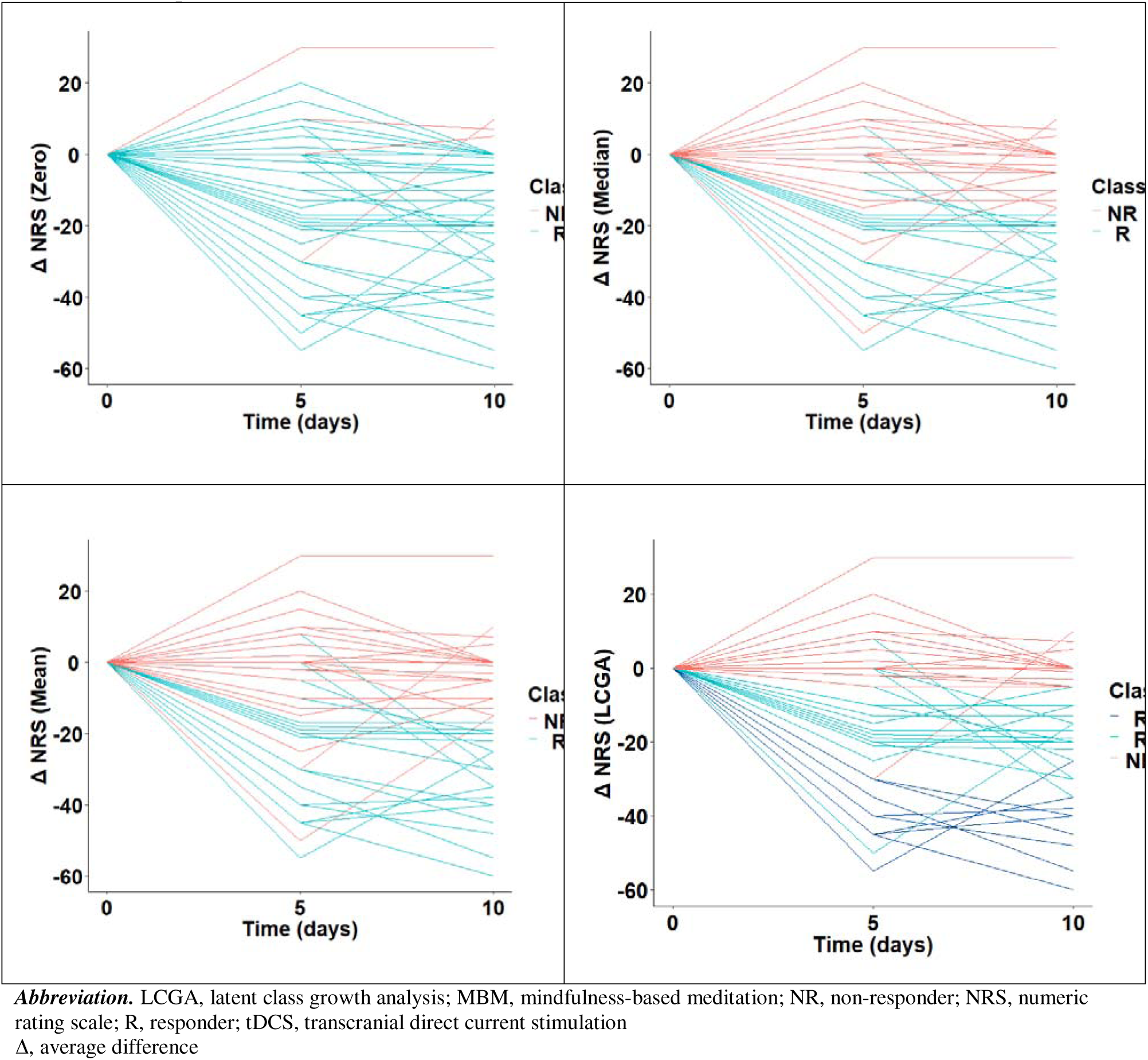
Group classification results for the first four methods: active tDCS + sham MBM.

**Figure 6.**
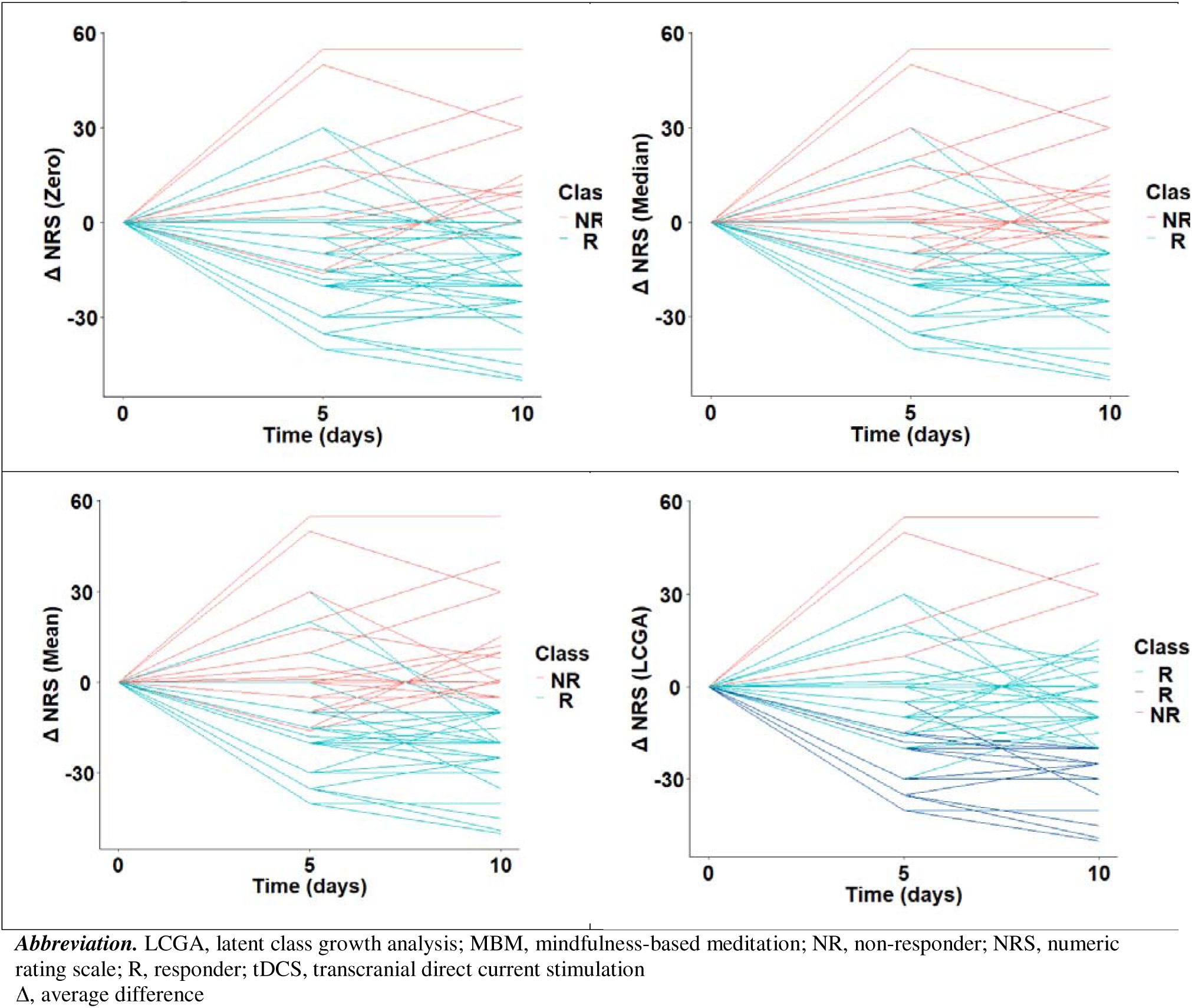
Group classification results for the first four methods: sham tDCS + active MBM.

**Figure 7.**
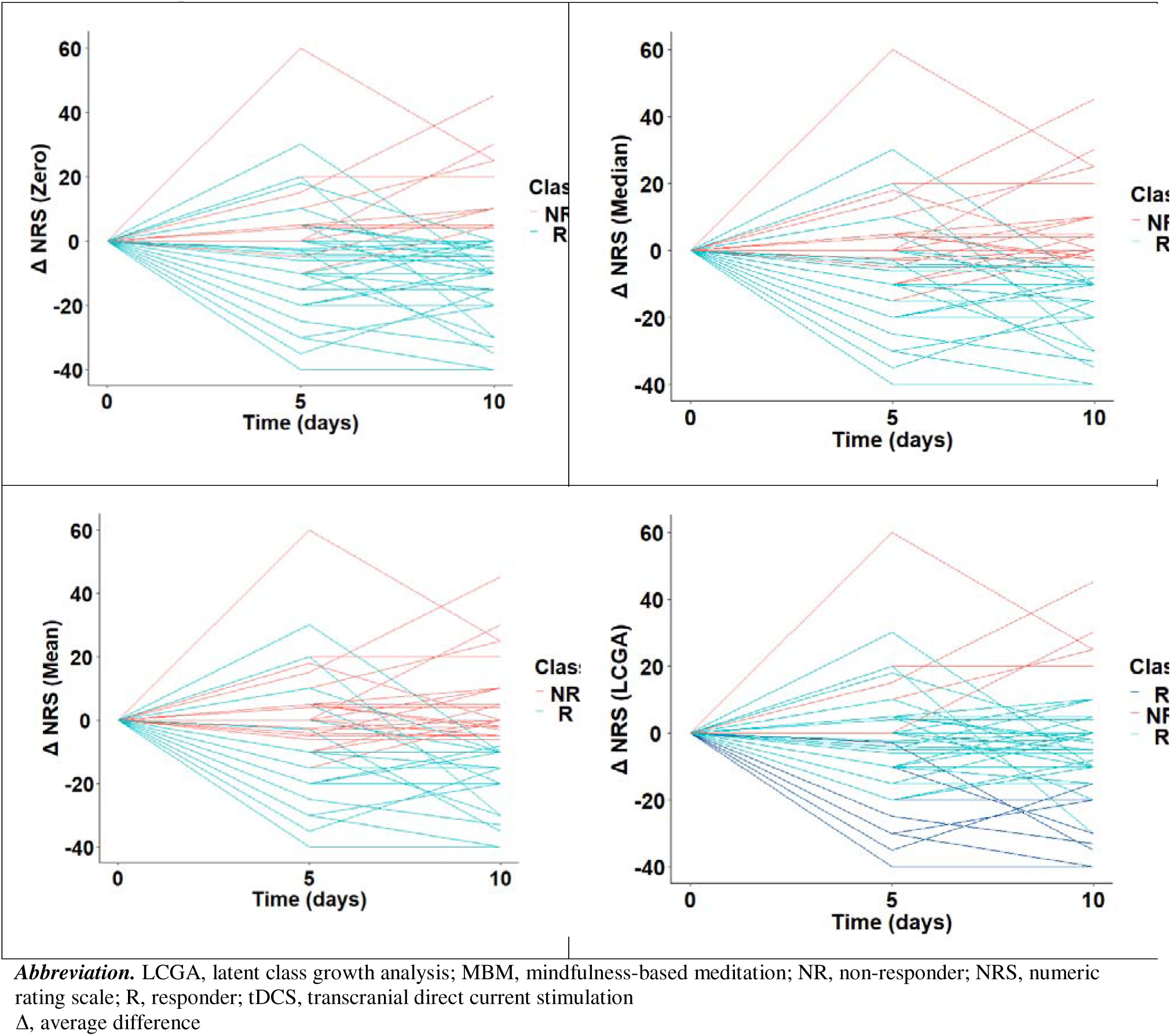
Group classification results for the first four methods: sham tDCS + sham MBM.

**Table 1.** Group classification results across four study groups.

| Method | Non-responder | Responder | Percentage<sup>a</sup> |
| --- | --- | --- | --- |
| active tDCS + active MBM |  |  |  |
| Zero | 5 | 45 | 90.0% |
| Median | 24 | 26 | 52.0% |
| Mean | 26 | 24 | 48.0% |
| LCGA | 30 | 20 | 40.0% |
| active tDCS + sham MBM |  |  |  |
| Zero | 4 | 46 | 92.0% |
| Median | 25 | 25 | 50.0% |
| Mean | 25 | 25 | 50.0% |
| LCGA | 17 | 33 | 66.0% |
| sham tDCS + active MBM |  |  |  |
| Zero | 13 | 37 | 74.0% |
| Median | 24 | 26 | 52.0% |
| Mean | 24 | 26 | 52.0% |
| LCGA | 5 | 45 | 90.0% |
| sham tDCS + sham MBM |  |  |  |
| Zero | 12 | 38 | 76.0% |
| Median | 23 | 27 | 54.0% |
| Mean | 27 | 23 | 46.0% |
| LCGA | 5 | 45 | 90.0% |

<sup>a</sup>This percentage refers to the proportion of the responders in the study group.
*Abbreviation.* tDCS, transcranial direct current stimulation; MBM, mindfulness-based meditation; LCGA, latent class growth analysis

**Table 2.**
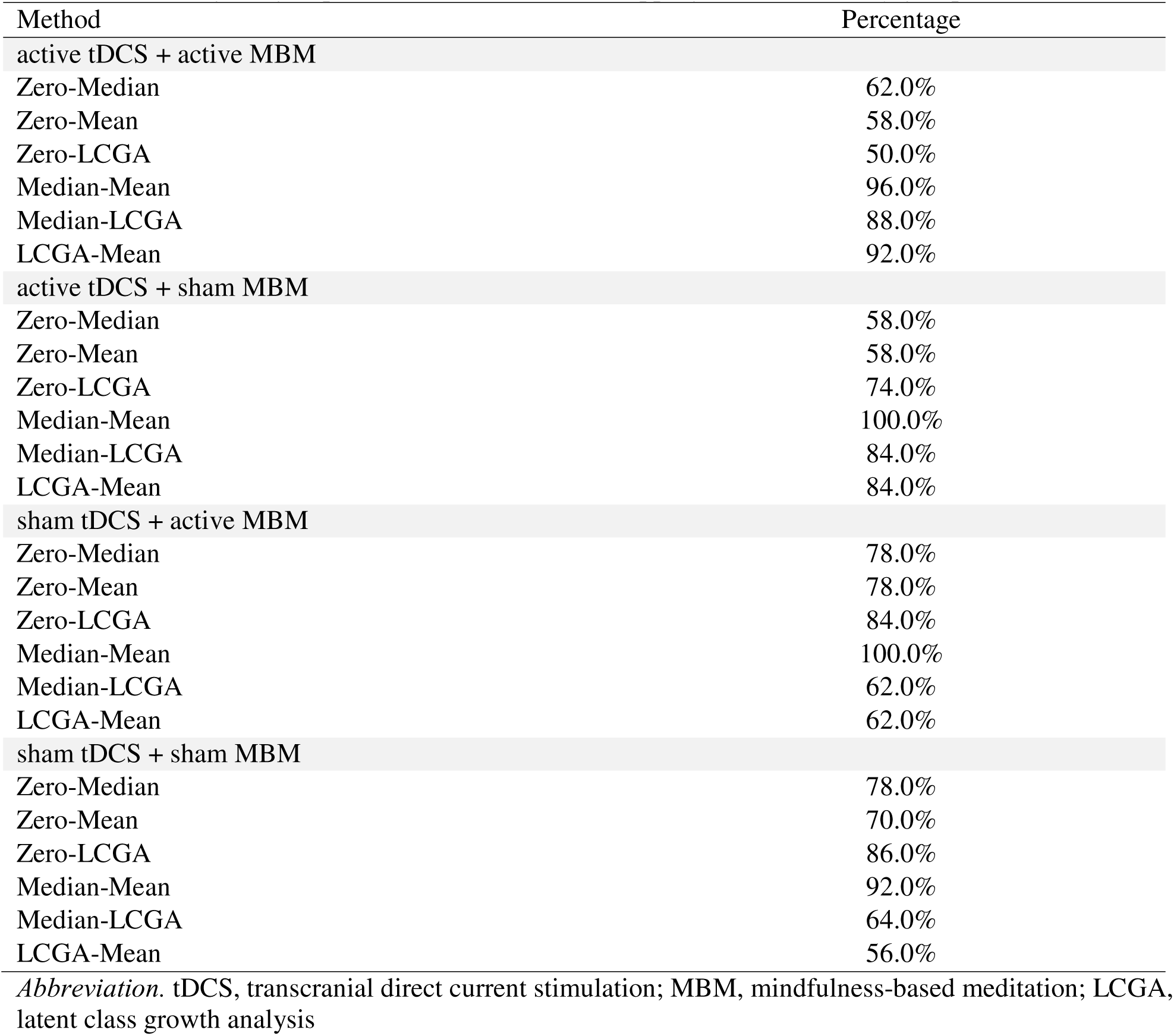
Percentage of group classification results overlapping across four study groups.

| Method | Percentage |
| --- | --- |
| active tDCS + active MBM |  |
| Zero-Median | 62.0% |
| Zero-Mean | 58.0% |
| Zero-LCGA | 50.0% |
| Median-Mean | 96.0% |
| Median-LCGA | 88.0% |
| LCGA-Mean | 92.0% |
| active tDCS + sham MBM |  |
| Zero-Median | 58.0% |
| Zero-Mean | 58.0% |
| Zero-LCGA | 74.0% |
| Median-Mean | 100.0% |
| Median-LCGA | 84.0% |
| LCGA-Mean | 84.0% |
| sham tDCS + active MBM |  |
| Zero-Median | 78.0% |
| Zero-Mean | 78.0% |
| Zero-LCGA | 84.0% |
| Median-Mean | 100.0% |
| Median-LCGA | 62.0% |
| LCGA-Mean | 62.0% |
| sham tDCS + sham MBM |  |
| Zero-Median | 78.0% |
| Zero-Mean | 70.0% |
| Zero-LCGA | 86.0% |
| Median-Mean | 92.0% |
| Median-LCGA | 64.0% |
| LCGA-Mean | 56.0% |

*Abbreviation.* tDCS, transcranial direct current stimulation; MBM, mindfulness-based meditation; LCGA, latent class growth analysis

### 3.1. Results by Study Groups

#### 3.1.1. active tDCS + active MBM

**Zero, mean, and median group criteria** Responders identified using these grouping criteria demonstrated greater improvements in clinical pain from baseline to post-intervention than non-responders (*p* < 0.001) (**Supplementary Table 2**). However, using zero as the classification criterion appeared oversimplified (**Figure 4**). Meanwhile, when classification was based on mean or median NRS scores, the non-responders and responders largely comprised the same members (**Table 2**).

**LCGA** LCGA produced clearer class separation than the other methods (**Figure 4**) and identified three classes. Two classes—weak and strong responders—were classified as responders and are represented by blue and navy lines, respectively. These responder classes showed a downward trajectory in the NRS change scores, whereas the non-responder class showed an upward trajectory. Overall, 40.0% of participants (*n* = 20) were classified as responders (**Table 1**) and demonstrated greater improvements in clinical pain from baseline to post-intervention than non-responders (*p* < 0.001; **Supplementary Table 2**).

Significant differences were observed in several baseline characteristics between responders and non-responders (**Supplementary Table 3**). Responders were younger (R: 64.05 [SD 7.80] vs. NR: 68.13 [SD 7.78]; *p* = 0.044), more likely to be female (R: 85.0% vs. NR: 56.7%; *p* = 0.035), and had a longer duration of osteoarthritis (R: 80.20 [SD 62.94] vs. NR: 51.57 [SD 55.44]; *p* = 0.043). In addition, responders exhibited significantly lower PPTh at the medial knee (R: 2.20 [SD 0.80] vs. NR: 3.35 [SD 1.47]; *p* = 0.004) and trapezius (R: 2.37 [SD 0.57] vs. NR: 3.58 [SD 1.95]; *p* = 0.009), as well as lower CPM (2.95 [SD 0.83] vs. 4.26 [SD 2.27]; *p* = 0.019). Moreover, β coefficients in response to punctate stimuli differed significantly between groups at the channel S06-D06 (R: 4.26 [SD 2.27] vs. NR: –0.97 [SD 4.11]; *p* = 0.003), channel S06-D08 (R: 1.50 [SD 2.62] vs. NR: –1.69 [SD 5.90]; *p* = 0.015), and the mPFC (R: 0.77 [SD 3.06] vs. NR: –1.12 [SD 3.32]; *p* = 0.017). β coefficients in response to heat stimuli also differed significantly at the channel S01-D03 (R: –1.01 [SD 1.76] vs. NR: 1.26 [SD 3.15]; *p* = 0.016), channel S04-D04 (R: 1.99 [SD 1.46] vs. NR: –0.50 [SD 2.36]; *p* = 0.006), and the rdlPFC (R: –1.21 [SD 2.66] vs. NR: 1.35 [SD 2.58]; *p* = 0.012).

A generalized linear model showed that β coefficients in response to punctate stimuli at S06-D06 (located in the lS1), race, and CPM significantly impacted the response group classification (*p* < 0.05); greater cortical activation at S06-D06, identifying as white, and lower CPM were significantly associated with being responders (**Table 3**).

**Table 3.** Generalized linear model analysis for LCGA classification results after stepwise BIC: active tDCS + active MBM.

| Variable | Estimate | Standard Error | 95% Confidence Intervals |  | *t* value | *p*-value |
| --- | --- | --- | --- | --- | --- | --- |
| *LCGA classification results<sup>a</sup>* |  |  |  |  |  |  |
| Intercept | -37.97 | 3131.80 | NA | 206.68 | -0.01 | 0.990 |
| BMI | 0.20 | 0.13 | -0.02 | 0.52 | 1.50 | 0.133 |
| Race, White | 32.15 | 3131.79 | -223.98 | NA | 0.01 | 0.992 |
| Average duration of osteoarthritis (months) | 0.02 | 0.01 | 0.00 | 0.06 | 1.71 | 0.088 |
| PPTh, medial knee | -1.08 | 0.81 | -3.05 | 0.24 | -1.33 | 0.183 |
| $\beta$ coefficients, S6D6 (punctate) | 1.07 | 0.52 | 0.29 | 2.40 | 2.04 | 0.042* |
| *LCGA classification results<sup>b</sup>* |  |  |  |  |  |  |
| Intercept | 7.72 | 6.17 | -3.84 | 21.38 | 1.25 | 0.211 |
| Age | -0.18 | 0.10 | -0.41 | 0.00 | -1.74 | 0.082 |
| BMI | 0.15 | 0.08 | 0.01 | 0.33 | 1.87 | 0.062 |
| Race, White | 4.84 | 2.35 | 1.02 | 10.54 | 2.06 | 0.039* |
| Average duration of osteoarthritis (months) | 0.01 | 0.01 | 0.00 | 0.03 | 1.64 | 0.101 |
| CPM | -1.78 | 0.87 | -3.86 | -0.44 | -2.04 | 0.041* |
| $\beta$ coefficients, IS1 (punctate) | 0.34 | 0.20 | 0.00 | 0.81 | 1.75 | 0.080 |

*Note.* Only variables that were significant in the univariate analysis (**Supplementary Table 3**) were entered into the model, while age, BMI, gender, and race were also included in the analysis even though they were not significant in the univariate tests. Some models did not converge and are not presented here.
$\beta$ coefficients from fNIRS channels and ROIs were modeled separately in the regression analysis to address multicollinearity. In addition, $\beta$ coefficients in response to punctate and heat stimuli were each modeled in separate regression models.
<sup>a</sup>Initial model included the $\beta$ coefficient from S06-D06 and S06-D08 in response to punctate stimuli.
<sup>b</sup>Initial model included the $\beta$ coefficient from IS1 in response to punctate stimuli.
*Abbreviations.* LCGA, latent class growth analysis; IS1, left somatosensory cortex; MBM, mindfulness-based meditation; mPFC, medial prefrontal cortex; PPTh, pressure pain threshold; tDCS, transcranial direct current stimulation.
Significance levels: \* $p < 0.05$

#### 3.1.2. active tDCS + sham MBM

**Zero, mean, and median group criteria** Responders identified using these grouping criteria demonstrated greater improvements in clinical pain from baseline to post-intervention compared to non-responders (*p* < 0.001) (**Supplementary Table 2**). Using zero as the classification criterion seemed oversimplified (**Figure 5**). Classifications based on the mean or median NRS scores yielded largely overlapping membership between responders and non-responders (**Table 2**).

**LCGA** LCGA again produced clearer class separation and identified three distinct classes (**Figure 5**). Non-responders, shown in red, exhibited an overall upward trajectory. Two additional classes—weak and strong responders—were classified as responder groups and are depicted in blue and navy; these classes generally showed a downward trend. Overall, 66.0% of participants (*n* = 33) were classified as responders (**Table 1**) and showed greater improvement in clinical pain from baseline to post-intervention than non-responders (*p* < 0.001) (**Supplementary Table 3**).

Responders showed significantly lower HPTh (R: 39.11 [SD 2.95] vs. NR: 41.88 [SD 4.27]; *p* = 0.022) and HPTo (R: 45.03 [SD 2.22] vs. NR: 46.63 [SD 2.72]; *p* = 0.020) at the knee compared to the non-responders (**Supplementary Table 4**). β coefficients in response to heat stimuli also differed significantly between groups at S08-D08 (R: –0.95 [SD 3.02] vs. NR: 1.38 [SD 3.45]; *p* = 0.047). According to the generalized linear model analysis, age and HPTo at the knee significantly impacted the response group classification (*p* < 0.05); younger age and lower HPTo at the knee were significantly associated with being responders (**Table 4**).

**Table 4.** Generalized linear model analysis for LCGA classification results after stepwise BIC: active t<u>DCS + sham MBM</u>

| Variable | Estimate | Standard Error | 95% Confidence Intervals |  | *t* value | *p*-value |
| --- | --- | --- | --- | --- | --- | --- |
| *LCGA classification results for NRS<sup>a</sup>* |  |  |  |  |  |  |
| Intercept | 38.66 | 14.93 | 13.21 | 73.81 | 2.59 | 0.010 |
| Age | -0.19 | 0.09 | -0.39 | -0.04 | -2.14 | 0.032* |
| HPT<sub>0</sub>, knee | -0.55 | 0.23 | -1.11 | -0.15 | -2.36 | 0.018* |

*Note.* Only variables that were significant in the univariate analysis (**Supplementary Tables 4**) were entered into the model, while age, body mass index, gender, and race were also included in the analysis even though they were not significant in the univariate tests. Some models did not converge and are not presented here.
$\beta$ coefficients from fNIRS channels and ROIs were modeled separately in the regression analysis to address multicollinearity. In addition, $\beta$ coefficients in response to punctate and heat stimuli were each modeled in separate regression models.
<sup>a</sup>Initial model included the $\beta$ coefficient from S08-D08 in response to heat stimuli
*Abbreviations.* LCGA, latent class growth analysis; MBM, mindfulness-based meditation; NRS, numeric rating scale; PPT<sub>h</sub>, pressure pain threshold; tDCS, transcranial direct current stimulation
Significance levels: \* $p < 0.05$

#### 3.1.3. sham tDCS + active MBM

Based on the group classification results from the first four methods (**Figure 6**), the individual trajectories did not form clear or distinct responder versus non-responder patterns. Instead, the trajectories were highly variable and often showed unstable directions. Some participants improved and then worsened, while others showed a brief increase by day 5 before declining, resulting in inconsistent classification boundaries between the two groups. Meanwhile, only five individuals were classified as non-responders in the LCGA; therefore, baseline comparisons were not conducted because of the substantial imbalance in group sizes.

#### 3.1.4. sham tDCS + sham MBM

The patterns across the first four classification methods were similar to those in the sham tDCS + active MBM group (**Figure 7**).

## 4. Discussion

The LCGA revealed the presence of HTE, identifying subgroups with differential responses (responders vs. non-responders), particularly in the combined tDCS and MBM group (active tDCS + active MBM) and the tDCS-only group (active tDCS + sham MBM). Several key baseline characteristics contributed to distinguishing responders from non-responders.

Notably, individuals with greater task-evoked cortical activation in channel S06-D06, located in the left S1, in response to punctate stimuli exhibited enhanced responsiveness to the combined intervention. Accumulating evidence suggests that clinically relevant interactions within and between pain-processing regions and higher-order networks influence the efficacy of analgesic treatments. For instance, pretreatment fMRI functional connectivity of pain-related brain regions has been shown to predict analgesic effects in individuals with fibromyalgia following anodal M1 tDCS^46^ and M1 repetitive transcranial magnetic stimulation.^47^ Jensen et al.^48^ also reported that between-person differences in response to neuromodulatory pain treatments were associated with baseline brain states, as indexed by EEG-assessed oscillatory activity. Extending this line of inquiry, our findings suggest that task-evoked cortical activation may serve as a neural marker for identifying subgroups of individuals with symptomatic KOA who differ in their responsiveness to a combined neuromodulatory and mindfulness-based intervention differs—a possibility that warrants further investigation. To date, fNIRS has primarily been used to characterize cortical activation changes following tDCS,^49^ including in individuals with KOA.^50,51^ However, studies examining fNIRS-derived cortical hemodynamic activity as a predictor of treatment response to non-pharmacological pain interventions remains limited. Replication of these findings in larger, well-powered samples will be essential to confirm their robustness and to advance the field toward a more mechanistic, biomarker-informed approach to pain management in KOA.

Individuals who identified as white demonstrated improved responsiveness to the combined tDCS and MBM intervention, which generally aligns with previous findings.^52,53^ For example, multidisciplinary pain treatment has been shown to be less effective in reducing self-reported pain severity among African American patients compared to white patients.^52^ Similarly, African American participants exhibited smaller improvements following a mindfulness-based integrative medical group visit intervention for chronic pain.^53^ Together, these findings suggest that ensuring equitable pain reduction across racial groups, whether through mind–body, cognitive-behavioral, or neuromodulatory therapies, remains an important priority.

Importantly, ongoing research is actively examining the mechanisms underlying observed differences in treatment response across racial groups. For instance, the PROACT trial protocol by Fillingim et al.^54^ is designed to evaluate whether a brief course of combined tDCS and mindfulness-based intervention produces comparable pain relief and central pain processing profiles among African American and white older adults with KOA, while also assessing biopsychosocial and neuroimaging markers that may explain racial differences in treatment response. Nonetheless, caution is warranted because our analysis collapsed all non-white participants into a single category, while white participants were disproportionately represented in our sample. This limitation parallels broader issues in the published literature on pain research. Although cognitive–behavioral therapies are well established as effective across chronic pain conditions, the evidence has been derived predominantly from white samples.^55^ Likewise, most current tDCS studies to date have been conducted primarily in white populations, highlighting the need for more inclusive and representative research to inform equitable pain management strategies.

This study found that younger individuals were significantly more likely to respond positively to tDCS. Chronological age has been identified as a relevant factor in contributing to the interindividual variability in the tDCS effect.^56^ This finding is supported by neurobiological mechanisms underlying tDCS, which depend on the propensity of an individual for plasticity induction. Indeed, such propensity tends to be higher at younger ages and declines across the lifespan, with a lower likelihood of occurring in older adulthood.^56,57^ Previous work has revealed that the capacity to induce plasticity via neuromodulation reduces with age.^58,59^ The decreased capacity for plasticity is, in part, reflected in older individuals through the diminished ability to generate long-term potentiation, which may limit the effectiveness of such neuromodulatory techniques.^60^ Furthermore, the reduction in brain volume associated with senescence increases the distance between the brain cortex and tDCS electrodes positioned over the scalp,^61^ which, in turn, can lead to a lower peak electric field under the tDCS electrodes in older individuals.^62^

CPM is commonly used to index the efficacy of descending pain pathways and can serve as an index of activation of endogenous pain inhibition.^63^ Yarnitsky et al.^64^ proposed using CPM as a tool to categorize patients by pain modulation profiles. Those with an inefficient CPM may exhibit a pronociception profile, which may predispose them to lower pain thresholds and a higher likelihood of developing chronic pain; in contrast, those with an efficient CPM may present with an antinociception profile and a higher pain threshold.^64^ This study found that individuals with a pronociceptive profile were significantly more likely to be responders to the combined tDCS and MBM intervention. Although not directly comparable, this finding echoes that of Yarnitsky et al.,^65^ who observed that medications acting on the descending pain modulation system were effective only in individuals with malfunctioning CPM, compared with those with high-functioning CPM in painful diabetic neuropathy. Thus, the combined intervention may have been effective primarily in those with inefficient CPM, as this intervention successfully engaged deficient inhibitory pathways, thereby contributing to reductions in clinical pain.

Those with lower HPTo at the knee were more likely to be classified as responders to tDCS, suggesting that heat pain sensitivity at the knee may play a role in forecasting treatment outcomes and underscoring the importance of assessing heat pain sensitivity for treatment selection. To our knowledge, the predictive value of baseline heat pain sensitivity has not been previously demonstrated. This may be because heat pain sensitivity is rarely assessed in treatment studies of chronic pain in KOA and is even less frequently examined as a potential predictor of treatment response, despite evidence indicating that improvements in heat pain sensitivity are associated with reductions in KOA pain.^66^ To date, only one study has reported evaluating HTE in response to tDCS, which found that heat pain sensitivity did not distinguish responders from non-responders.^67^ However, a direct comparison with our findings is limited by methodological differences in how the treatment response was defined. Indeed, the prior study used a multi-trajectory modeling approach to identify latent subgroups based on jointly modeled trajectories of knee pain and symptom severity (i.e., NRS scores and the Western Ontario and McMaster Universities Osteoarthritis Index) from baseline to three months post-intervention.^67^ In contrast, the present study identified subgroups using change scores (i.e., change in NRS) from baseline to the immediate post-10-day intervention period.

In contrast to the tDCS-containing arms, the MBM-only group (sham tDCS + active MBM) showed markedly smaller changes in pain intensity over the 10-day intervention. MBM engages attentional and emotional regulation pathways, which can reduce pain perception and improve coping strategies;^68–70^ however, these effects may not manifest as immediate reductions in pain intensity, particularly within a short treatment window. Moreover, the benefits of meditation may wane without continued practice.^71^ Therefore, the magnitude of MBM-related improvement may have been too subtle within this brief interval to generate distinct response patterns, thereby limiting our ability to identify clear responder profiles. A similar rationale may apply to the sham group (sham tDCS + sham MBM). The small magnitude of change in pain intensity in this group was likely reflective of a sham effect, suggesting that a sham protocol previously considered inactive may still exert minor neuromodulatory effects on certain outcomes.^72^ Prior studies have indicated that such effects may arise from skin sensations initially induced in the sham condition (e.g., ramp-up/down procedures) or from cortical modulation by microampere-scale currents.^73,74^

Typically, affective interference leads to lower adherence to treatment protocols. One systematic review highlighted the influence of psychological factors on treatment response across multiple modalities (e.g., pharmacological, physical therapy, and combined treatments),^75^ showing that higher levels of depressive symptoms, anxiety, and pain catastrophizing reduced the likelihood of treatment success.^76^ Teixeira et al.^25^ found that lower scores on psychological functioning measures—the SF-12 Mental Component Summary and the Beck depression inventory—were the most important predictors of an analgesic response to tDCS in patients with chronic KOA pain and a dysfunctional descending pain inhibitory system. Kambeitz et al.^26^ found that negative effects and the number of depressive episodes strongly predicted the treatment response for prefrontal tDCS in patients with major depression.

Interestingly, this study found that baseline psychological characteristics did not distinguish responders from non-responders in either the combined tDCS + MBM group or the tDCS-only group. Likewise, in the study by Gunduz et al.^27^ involving patients with phantom limb pain receiving M1–tDCS, psychological factors were not identified as meaningful predictors of treatment response. The factors underlying HTE in neuromodulation may vary across treatment protocols, neural targets, and population characteristics. Nonetheless, psychological factors may still function as effect modifiers or mediators in specific contexts, underscoring the need for further investigation to inform optimized and targeted intervention design.

### 4.1. Limitations

Some limitations should be acknowledged. First, the current analyses were exploratory and not prespecified in the trial, meaning cautious interpretation of the conclusions is required. Second, the generalizability of the study to both sexes could also be negatively affected by the large proportion of females in our sample. Third, although this was the largest trial to date examining tDCS and MBM treatment in individuals with KOA, the sample size may still be insufficient to draw robust conclusions in this HTE analysis, which relies on subgroup comparisons. Fourth, our study relied on individual participant characteristics examined in isolation, whereas complex interactions among factors are likely to influence treatment response. Understanding these interactions in future research will help advance the field toward a personalized, mechanism-based treatment approach for KOA. For instance, one study demonstrated that combining preoperative CPM with pain catastrophizing produced a stronger predictive model for treatment response to total knee arthroplasty than either assessment alone.^77^ Finally, the analysis of our fNIRS data primarily relied on statistical analysis centered on β values, focusing on spatial activation patterns. While this approach provides valuable insights, future research could benefit from incorporating time domain analysis techniques to delve deeper into cortical activation patterns.

## 5. Conclusion

Evaluating the HTE is essential for identifying which subgroups benefit most from an intervention and under what conditions it is most effective. In this study of individuals with symptomatic KOA, several factors were associated with HTE in the combined tDCS and MBM intervention, providing preliminary insights that may inform the development of personalized pain management strategies. Thus, future tDCS trials should also consider the characteristics underlying the observed HTE when designing interventions and selecting participants.

## Supporting information

Clean Manuscript

## Declaration Statements

### Data Availability

All data supporting the findings of this study are summarized in this published article. Requests for additional de-identified data will be reviewed by the corresponding author on a case-by-case basis and may require institutional and ethical approval.

## Acknowledgements

We thank Dr. Alejandro Chaoul for his valuable contributions as a consultant on the meditation component of this project. His expertise in mind–body practices informed the development and implementation of the intervention. This work was supported by the National Institute of Nursing Research (R01 NR019051-01, 2020–2025; R01 NR019051-03S1, 2022–2025), the National Institute on Aging (R03 AG093555-01, 2025–2027), and the University of Arizona College of Nursing.

