## Supplementary material for "Neural and psychophysical predictors of treatment response to transcranial direct current stimulation and mindfulness-based meditation for knee osteoarthritis pain": Clean Manuscript

### **Supplementary Material S1.** Quantitative sensory testing (QST) procedures

**Thermal Testing Procedures.** Contact heat stimuli were delivered using a computer-controlled TSA-II NeuroSensory Analyzer (Medoc Ltd., Ramat Yishai, Israel) to assess heat pain threshold (HPTh) and heat pain tolerance (HPTo) at both the index knee and the ipsilateral ventral forearm, employing an ascending method of limits. To prevent sensitization or habituation of cutaneous receptors, the thermode position was rotated among three locations between trials at each body site. Starting at a baseline temperature of 32°C, the thermode temperature increased at a rate of 0.5°C per second until participants pressed a button on a handheld device. Participants were instructed to press the button when they first perceived the heat as painful to measure HPTh and when the heat became intolerable to assess HPTo. Three HPTh trials were conducted at the first test site, followed by three HPTo trials. The same sequence was then repeated at the second test site, with a 5-minute rest period between sites. The average of the three trials was calculated for each participant to determine overall HPTh and HPTo temperatures for analysis.

**Mechanical Testing Procedures.** Pressure pain threshold (PPTh) was evaluated using a handheld digital pressure algometer (Wagner, Greenwich, Connecticut, USA) to apply blunt mechanical pressure to deep tissues, including muscles and joints. The pressure was increased at a constant rate of 0.3 kgf/cm² per second to determine PPTh at two locations: the medial side of the index knee and the trapezius muscle. The order of testing sites was randomized and counterbalanced. Participants were instructed to indicate the point at which the sensation “first became painful” and at which point the applied pressure was recorded. The average of three trials at each site was calculated to determine the PPTh for that location.

After assessing PPTh, participants underwent testing to evaluate cutaneous mechanical sensitivity to punctate stimuli on both the index patella and back of the ipsilateral hand. A calibrated nylon monofilament delivering a target force of 300 g was applied 10 times at a rate of one contact per second, with participants providing verbal pain intensity ratings on a scale from 0 (no pain sensation) to 100 (the most intense pain imaginable). Pain ratings from the two trials were averaged to punctate mechanical pain at each site. For TSP assessment, participants first rated the pain intensity from a single application of the monofilament. They then rated the maximum pain intensity experienced during a series of 10 contacts administered at a rate of one contact per second. Temporal summation of pain was calculated by subtracting the pain rating for the single stimulus from the rating for the series of 10 stimuli at each site.

**CPM*.*** Ten minutes after assessing thermal or mechanical pain, CPM was evaluated by calculating the change in PPTh during the cold pressor task relative to baseline PPTh. Baseline PPTh measurements were obtained immediately before participants immersed their hands in a cold water bath maintained at 12°C. Thirty seconds after immersion, participants rated their cold pain intensity on a scale from 0 to 100, followed by a second PPTh measurement. Participants were instructed to keep their hands submerged for as long as tolerable for up to a maximum of one minute. Upon hand removal, the final PPTh measurements were recorded. CPM at 60 seconds was calculated by subtracting baseline PPTh from PPTh at 60 seconds. The 12°C temperature was selected based on prior studies involving middle-aged and older adults with KOA, as it was found to induce moderate yet tolerable pain in most participants (King et al., 2013). The water was continuously circulated and maintained at a stable temperature using a refrigeration unit (Neslab, Portsmouth, New Hampshire, USA). An increase in PPTh following cold water immersion indicated pain inhibition.

The sequence of heat and mechanical testing was randomized and counterbalanced, with CPM always administered last to minimize any potential carryover effects. The same researcher performed QST on each participant throughout the study, and all participants received standardized recorded instructions to prevent bias during data collection and enhance the reliability of the results.

### **Supplementary Table 1.** Baseline characteristics of participants by study group

|  | *n* (%) or mean ± standard deviation | | | | |  |
| --- | --- | --- | --- | --- | --- | --- |
|  | active tDCS + active MBM  (*n* = 53) | active tDCS + sham MBM  (*n* = 50) | sham tDCS + active MBM  (*n* = 53) | sham tDCS + sham MBM  (*n* = 52) | Total  (N = 208) | *p*-value |
| Age, years | 66.49 (7.74) | 68.06 (7.42) | 67.19 (7.89) | 68.88 (6.92) | 67.64 (7.51) | 0.590 |
| Body mass index (kg/m^2^) | 31.83 (8.72) | 29.03 (6.95) | 31.30 (8.41) | 31.16 (7.69) | 30.85 (8.00) | 0.347 |
| Gender |  |  |  |  |  | 0.597 |
| male | 19 (35.8%) | 16 (32.0%) | 13 (24.5%) | 18 (34.6%) | 66 (31.7%) |  |
| female | 34 (64.2%) | 34 (68.0%) | 40 (75.5%) | 34 (65.4%) | 142 (68.3%) |  |
| Race |  |  |  |  |  | 0.711 |
| non-white | 9 (17.0%) | 7 (14.0%) | 12 (22.6%) | 9 (17.3%) | 37 (17.8%) |  |
| white | 44 (83.0%) | 43 (86.0%) | 41 (77.4%) | 43 (82.7%) | 171 (82.2%) |  |
| Education |  |  |  |  |  | 0.960 |
| high school or less | 11 (20.8%) | 10 (20.0%) | 10 (18.9%) | 12 (23.1%) | 43 (20.7%) |  |
| college or higher | 42 (79.2%) | 40 (80.0%) | 43 (81.1%) | 40 (76.9%) | 165 (79.3%) |  |
| Marital status |  |  |  |  |  | 0.201 |
| married/partnered | 35 (66.0%) | 37 (74.0%) | 29 (54.7%) | 31 (59.6%) | 132 (63.5%) |  |
| unmarried/unpartnered | 18 (34.0%) | 13 (26.0%) | 24 (45.3%) | 21 (40.4%) | 76 (36.5%) |  |
| Average duration of osteoarthritis (months) | 62.49 (59.01) | 34.86 (39.97) | 59.42 (71.00) | 50.92 (58.58) | 52.17 (58.98) | 0.019* |
| Kellgren-Lawrence score (index knee) |  |  |  |  |  | 0.954 |
| 0-1 | 7 (13.2%) | 6 (12.0%) | 8 (15.1%) | 8 (15.4%) | 29 (13.9%) |  |
| ≥ 2 | 46 (86.8%) | 44 (88.0%) | 45 (84.9%) | 44 (84.6%) | 179 (86.1%) |  |
| NRS.0^a^ | 42.40 (25.79) | 44.94 (25.33) | 41.83 (21.98) | 42.13 (24.17) | 42.80 (24.20) | 0.886 |
| NRS.5^b^ | 29.46 (20.83) | 31.56 (26.32) | 36.72 (22.36) | 38.59 (28.10) | 34.08 (24.67) | 0.167 |
| NRS.10^c^ | 23.97 (16.80) | 28.16 (25.09) | 35.32 (22.84) | 36.42 (27.94) | 30.97 (23.91) | 0.046* |
| Difference in NRS^d^ | -19.41 (19.76) | -16.78 (17.90) | -7.16 (22.24) | -6.20 (17.99) | -12.39 (20.26) | 0.003* |
| HPTh, knee (°C) | 40.92 (3.31) | 40.05 (3.66) | 40.33 (3.28) | 41.62 (3.29) | 40.74 (3.41) | 0.079 |
| HPTo, knee (°C) | 46.01 (2.24) | 45.57 (2.50) | 45.33 (2.49) | 45.55 (2.67) | 45.62 (2.47) | 0.535 |
| HPTh, arm (°C) | 39.82 (3.54) | 38.02 (2.79) | 38.09 (3.07) | 38.52 (3.15) | 38.62 (3.22) | 0.021* |
| HPTo, arm (°C) | 45.58 (2.70) | 44.35 (3.48) | 44.13 (3.10) | 44.85 (3.35) | 44.73 (3.19) | 0.067 |
| PPTh, medial knee (kgf) | 2.88 (1.32) | 2.72 (1.45) | 2.70 (1.34) | 2.76 (1.24) | 2.76 (1.33) | 0.779 |
| PPTh, trapezius (kgf) | 3.07 (1.61) | 2.73 (1.38) | 2.78 (1.08) | 2.83 (1.25) | 2.85 (1.34) | 0.635 |
| Punctate mechanical pain, patella (average series of 10 in patella punctate mechanical, score: 0-100) | 8.02 (13.07) | 8.72 (11.92) | 10.66 (16.18) | 13.07 (19.82) | 10.12 (15.59) | 0.666 |
| Temporal summation of pain, patella (series of 10 - single trial) | 3.26 (11.17) | 3.52 (9.21) | 4.13 (10.62) | 6.04 (15.62) | 4.24 (11.88) | 0.809 |
| Punctate mechanical pain, hand (average series of 10 in patella punctate mechanical, score: 0-100) | 4.73 (7.99) | 8.59 (14.38) | 5.71 (9.95) | 10.45 (14.74) | 7.34 (12.19) | 0.363 |
| Temporal summation of pain, hand (series of 10 - single trial) | 2.74 (6.38) | 3.41 (8.80) | 2.94 (8.11) | 4.58 (8.91) | 3.41 (8.07) | 0.916 |
| CPM (PPTh at 60 seconds - PPTh at pre-CPM) | 3.70 (1.89) | 3.13 (1.31) | 3.24 (1.28) | 3.26 (1.47) | 3.34 (1.52) | 0.449 |
| Cold pain intensity (score: 0-100) | 72.13 (27.12) | 68.30 (29.60) | 74.43 (24.49) | 72.31 (26.13) | 71.84 (26.76) | 0.844 |
| Depression (CES-D) | 16.96 (5.13) | 18.90 (8.08) | 17.70 (6.13) | 18.58 (6.08) | 18.02 (6.42) | 0.539 |
| Pain catastrophizing (PCS) | 9.00 (7.84) | 12.08 (10.35) | 10.66 (9.04) | 10.42 (8.26) | 10.52 (8.91) | 0.457 |
| COVID stress (CSS) | 9.51 (11.08) | 10.76 (10.89) | 10.28 (13.67) | 12.62 (11.59) | 10.78 (11.84) | 0.419 |
| *β coefficients (HbO) by fNIRS channel in response to punctate stimuli* |  |  |  |  |  |  |
| S01-D01 | -0.06 (3.47) | 0.40 (3.65) | -0.32 (4.43) | 0.01 (4.36) | -0.00 (3.95) | 0.985 |
| S01-D03 | -0.90 (2.59) | 0.27 (2.50) | 0.58 (3.22) | 0.35 (3.62) | 0.07 (3.04) | 0.288 |
| S02-D02 | 0.77 (3.46) | -0.95 (4.62) | -0.42 (4.23) | 1.80 (2.71) | 0.43 (3.83) | 0.057 |
| S02-D04 | 1.23 (4.18) | -1.42 (5.45) | -1.36 (7.79) | 0.35 (3.00) | -0.22 (5.36) | 0.399 |
| S03-D01 | -0.03 (2.72) | -1.30 (2.69) | 0.23 (3.80) | -0.92 (5.24) | -0.50 (3.88) | 0.299 |
| S03-D03 | 0.34 (4.14) | -0.32 (3.01) | 0.40 (3.22) | 0.11 (3.63) | 0.15 (3.51) | 0.866 |
| S03-D05 | -0.16 (3.39) | 0.04 (3.21) | 0.33 (3.95) | -0.35 (3.25) | -0.05 (3.41) | 0.727 |
| S04-D02 | -0.65 (4.24) | 0.03 (4.47) | -1.06 (6.75) | -1.87 (5.72) | -0.90 (5.36) | 0.591 |
| S04-D04 | -0.05 (3.11) | -0.10 (2.33) | 0.70 (3.05) | 0.09 (3.94) | 0.14 (3.19) | 0.738 |
| S04-D06 | 0.29 (2.78) | -0.11 (2.75) | 0.27 (2.72) | 0.27 (4.42) | 0.20 (3.33) | 0.899 |
| S05-D03 | 1.89 (3.60) | -0.17 (4.24) | 0.73 (3.03) | 0.42 (3.35) | 0.73 (3.58) | 0.411 |
| S05-D05 | 0.31 (4.45) | -0.88 (6.32) | 1.02 (3.48) | -0.16 (5.18) | 0.05 (4.96) | 0.777 |
| S05-D07 | 0.93 (2.85) | -1.20 (3.32) | -0.50 (3.27) | 0.66 (3.79) | 0.03 (3.40) | 0.147 |
| S06-D04 | 0.98 (3.28) | -1.47 (5.72) | 1.10 (6.31) | 0.20 (5.46) | 0.27 (5.29) | 0.542 |
| S06-D06 | -0.03 (3.53) | 0.55 (4.10) | -0.53 (2.89) | -0.33 (3.41) | -0.09 (3.49) | 0.862 |
| S06-D08 | -0.31 (4.97) | 1.09 (4.32) | -0.36 (3.74) | -0.38 (4.60) | 0.02 (4.46) | 0.794 |
| S07-D05 | 0.46 (4.47) | 1.38 (4.14) | 0.19 (2.76) | -0.19 (3.82) | 0.37 (3.83) | 0.861 |
| S07-D07 | -0.17 (3.93) | 0.33 (3.12) | 0.01 (2.71) | 0.15 (4.73) | 0.08 (3.77) | 0.755 |
| S08-D06 | -1.30 (4.97) | 0.14 (3.84) | 0.47 (4.41) | 0.05 (3.43) | -0.17 (4.17) | 0.567 |
| S08-D08 | -0.66 (3.62) | -0.17 (4.02) | -0.42 (2.87) | 0.30 (4.24) | -0.21 (3.74) | 0.729 |
| *β coefficients (HbO) by fNIRS ROIs in response to punctate stimuli* |  |  |  |  |  |  |
| Right dorsolateral prefrontal cortex (rdlPFC) | -0.33 (2.57) | -0.15 (2.67) | 0.38 (2.80) | -0.10 (3.08) | -0.06 (2.78) | 0.845 |
| Medial prefrontal cortex (mPFC) | 0.37 (1.90) | -0.72 (2.90) | 0.73 (2.00) | -0.08 (2.40) | 0.06 (2.37) | 0.158 |
| Left dorsolateral prefrontal cortex (ldlPFC) | 0.40 (2.90) | 0.21 (2.34) | 0.17 (2.46) | 0.27 (2.52) | 0.27 (2.55) | 0.789 |
| Right somatosensory cortex (rS1) | 0.02 (3.31) | -0.60 (2.77) | -0.58 (3.80) | -0.25 (2.42) | -0.34 (3.07) | 0.705 |
| Left somatosensory cortex (lS1) | -0.34 (3.31) | 0.29 (2.59) | -0.22 (2.79) | -0.11 (2.46) | -0.09 (2.79) | 0.616 |
| *β coefficients (HbO) by fNIRS channel in response to heat stimuli* |  |  |  |  |  |  |
| S01-D01 | 1.48 (5.57) | -0.65 (3.24) | 0.07 (4.04) | -0.28 (4.22) | 0.15 (4.38) | 0.250 |
| S01-D03 | 0.48 (2.93) | -0.16 (2.41) | 0.09 (4.19) | 0.05 (4.30) | 0.11 (3.50) | 0.700 |
| S02-D02 | 0.67 (3.27) | 1.89 (3.37) | -0.04 (2.90) | -0.32 (4.27) | 0.47 (3.61) | 0.076 |
| S02-D04 | 0.61 (4.57) | 1.66 (3.30) | 0.59 (2.83) | -0.31 (5.20) | 0.56 (4.22) | 0.060 |
| S03-D01 | -0.38 (3.35) | 0.22 (2.96) | -0.60 (3.45) | 1.08 (5.00) | 0.14 (3.88) | 0.991 |
| S03-D03 | 1.17 (4.02) | -0.06 (3.22) | -0.20 (4.82) | 1.20 (5.54) | 0.57 (4.56) | 0.637 |
| S03-D05 | -0.86 (2.71) | -0.09 (2.62) | -0.12 (2.85) | -0.39 (3.16) | -0.37 (2.83) | 0.491 |
| S04-D02 | -0.90 (3.99) | -0.85 (3.72) | 0.41 (3.64) | 0.47 (4.58) | -0.23 (4.03) | 0.299 |
| S04-D04 | 0.45 (2.37) | 1.15 (3.34) | 0.86 (3.03) | -0.62 (3.21) | 0.38 (3.03) | 0.507 |
| S04-D06 | 0.69 (3.20) | 1.13 (2.55) | 0.08 (3.82) | -0.57 (3.79) | 0.27 (3.43) | 0.151 |
| S05-D03 | 0.72 (3.68) | 0.76 (2.36) | 0.90 (4.42) | -0.82 (2.94) | 0.25 (3.36) | 0.033 |
| S05-D05 | 0.89 (5.10) | -0.39 (5.79) | 1.26 (6.02) | -0.92 (4.38) | 0.09 (5.28) | 0.370 |
| S05-D07 | -1.32 (2.64) | -1.86 (6.04) | -0.61 (4.59) | -0.39 (4.14) | -1.01 (4.43) | 0.511 |
| S06-D04 | -0.25 (4.76) | -3.72 (8.30) | -1.40 (6.41) | -0.92 (5.92) | -1.50 (6.45) | 0.257 |
| S06-D06 | -1.70 (4.83) | 0.44 (2.10) | -0.50 (3.33) | -0.05 (2.63) | -0.46 (3.44) | 0.103 |
| S06-D08 | -1.74 (6.84) | -0.13 (3.09) | -0.99 (4.03) | 1.18 (5.64) | -0.36 (5.22) | 0.222 |
| S07-D05 | 0.51 (3.24) | 0.81 (2.27) | -0.27 (1.98) | -0.36 (2.21) | 0.14 (2.50) | 0.408 |
| S07-D07 | 0.90 (3.89) | 0.82 (2.67) | -0.02 (3.08) | -0.18 (2.65) | 0.35 (3.11) | 0.599 |
| S08-D06 | -0.06 (3.81) | 0.06 (2.92) | -0.63 (3.30) | 0.33 (5.06) | -0.06 (3.92) | 0.561 |
| S08-D08 | 0.17 (3.65) | 0.06 (3.36) | -0.71 (3.81) | -0.35 (5.62) | -0.21 (4.28) | 0.619 |
| *β coefficients (HbO) by fNIRS ROIs in response to heat stimuli* |  |  |  |  |  |  |
| Right dorsolateral prefrontal cortex (rdlPFC) | 0.32 (2.87) | -0.21 (1.92) | 0.18 (3.02) | 0.41 (3.08) | 0.18 (2.74) | 0.740 |
| Medial prefrontal cortex (mPFC) | 0.25 (2.84) | -0.43 (4.34) | 0.21 (2.82) | -0.23 (2.44) | -0.07 (3.17) | 0.868 |
| Left dorsolateral prefrontal cortex (ldlPFC) | 0.04 (2.69) | 0.22 (3.26) | -0.21 (3.13) | -0.49 (1.91) | -0.14 (2.71) | 0.686 |
| Right somatosensory cortex (rS1) | -0.27 (3.01) | 0.47 (2.87) | 0.50 (2.94) | -0.29 (2.99) | 0.08 (2.95) | 0.284 |
| Left somatosensory cortex (lS1) | -0.71 (3.97) | 0.02 (2.16) | -0.65 (2.66) | 0.31 (3.15) | -0.24 (3.08) | 0.437 |

*Abbreviation.* CES-D, Center for Epidemiologic Studies Depression Scale; CPM, conditioned pain modulation; CSS, COVID stress scales; PCS, pain catastrophizing scale; PPTh, pressure pain threshold; HPTh, heat pain threshold, HPTo, heat pain tolerance; tDCS, transcranial direct current stimulation; MBM, mindfulness-based meditation; NRS, numerical rating scale; ROI, region of interest; HbO, oxygenated hemoglobin concentration

^a^NRS average pain in the past 24 hours at baseline

^b^NRS average pain in the past 24 hours on day 5 of intervention

^c^NRS average pain in the past 24 hours on day 10 of intervention

^d^NRS.10-NRS.0

Significance levels: **p* < 0.05

**Supplementary Figure 1.** Group classification results for GBMTM

| 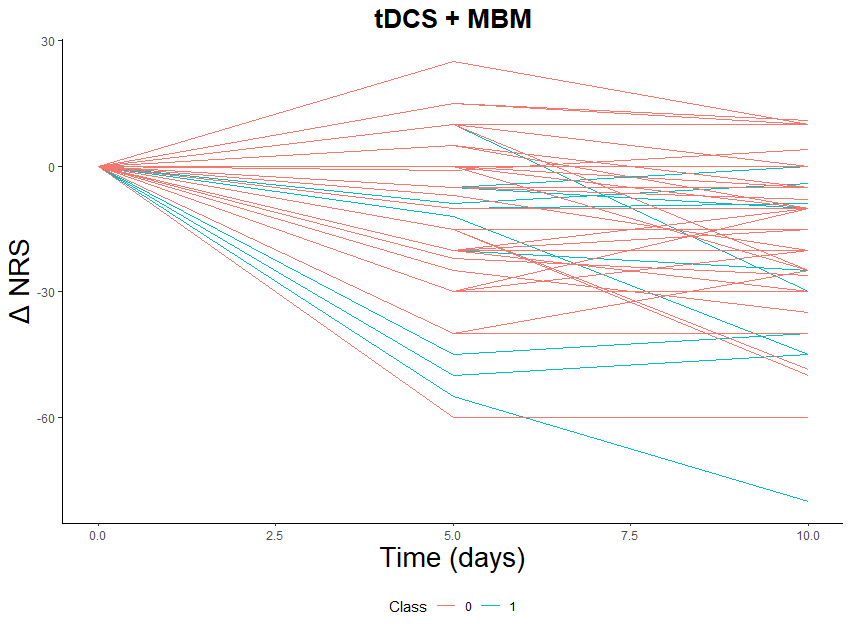 | 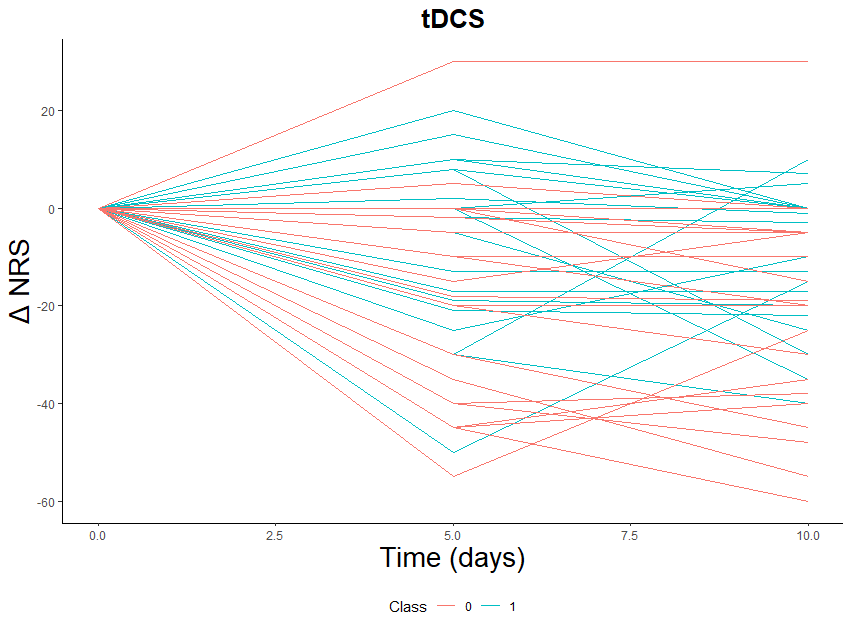 |
| --- | --- |
| 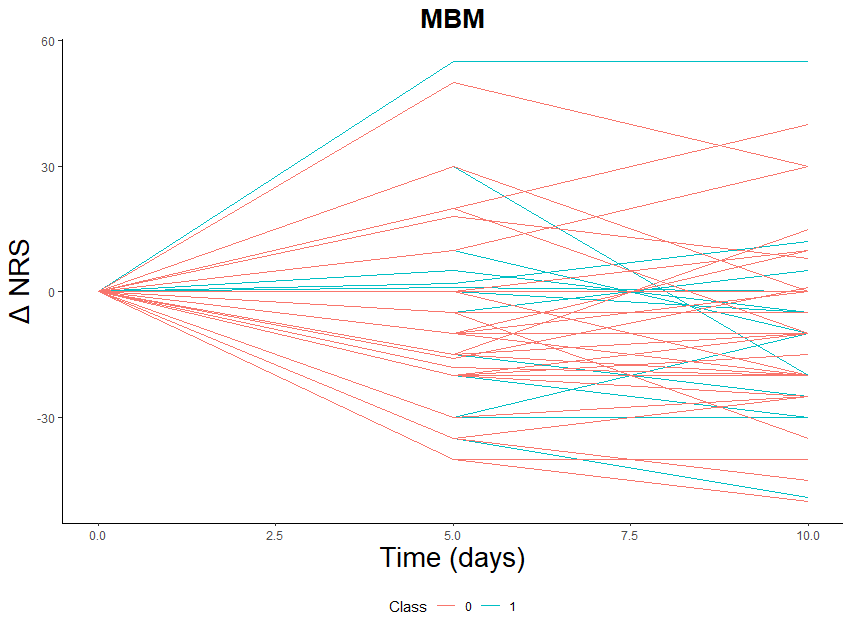 | 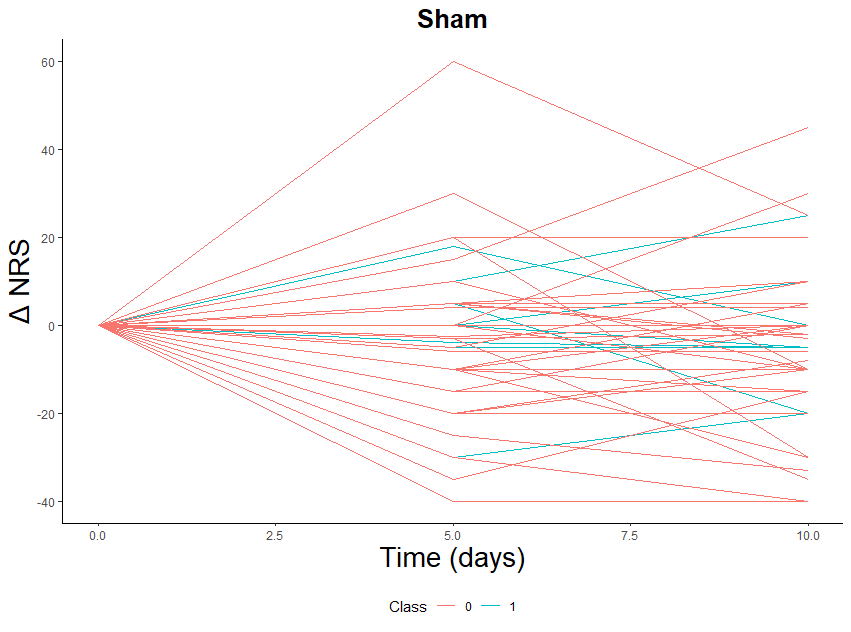 |

*Abbreviation.* MBM, mindfulness-based meditation; NRS, Numeric Rating Scale; tDCS, transcranial direct current stimulation

Δ, average difference

### **Supplementary Table 2.** Comparison between two groups on changes from baseline in treatment outcome (NRS): active tDCS + active MBM

| Variable | Non-responder | Responder | Effect size | *p*-value |
| --- | --- | --- | --- | --- |
| active tDCS + active MBM |  |  |  |  |
| Average difference—Zero^b^ | 9.00 (2.83) | -22.57 (18.2) | 1.81 | < 0.001 |
| Average difference—Median^b^ | -2.96 (7.26) | -34.60 (14.7) | 2.69 | < 0.001 |
| Average difference—Mean^b^ | -3.88 (7.70) | -36.23 (14.10) | 2.87 | < 0.001 |
| Average difference—LCGA^b^ | -6.70 (10.3) | -38.48 (14.4) | 2.63 | < 0.001 |
| active tDCS + sham MBM |  |  |  |  |
| Average difference—Zero^a^ | 13.00 (11.5) | -19.37 (16.0) | 2.06 | < 0.001 |
| Average difference—Median^b^ | -2.60 (9.32) | -30.96 (12.2) | 2.61 | < 0.001 |
| Average difference—Mean^b^ | -2.60 (9.32) | -30.96 (12.2) | 2.61 | < 0.001 |
| Average difference —LCGA^b^ | 1.35 (8.58) | -26.12 (13.8) | 2.24 | < 0.001 |

^a^ Variables analyzed by t Test

^b^ Variables analyzed by Wilcoxon rank-sum test.

*Note.* Depending on data distribution, the Wilcoxon rank-sum test was used for score changes with non-normal distributions, and the t test was used for score changes with normal distributions.

*Abbreviation.* MBM, mindfulness-based meditation; NRS, numeric rating scale; tDCS, transcranial direct current stimulation

### **Supplemental Table 3.** LCGA classification results for NRS: active tDCS + active MBM

|  | *n* (%) or mean ± standard deviation | | |  |
| --- | --- | --- | --- | --- |
|  | Non-responder (*n* = 30) | Responder (*n* = 20) | Total (*n* = 50) | *p*-value |
| Age, years | 68.13 (7.78) | 64.05 (7.80) | 66.50 (7.97) | 0.044* |
| Body mass index (kg/m^2^) | 31.62 (9.87) | 32.31 (7.18) | 31.90 (8.82) |  |
| Gender |  |  |  | 0.035* |
| male | 13 (43.3%) | 3 (15.0%) | 16 (32.0%) |  |
| female | 17 (56.7%) | 17 (85.0%) | 34 (68.0%) |  |
| Race |  |  |  | 0.229 |
| non-white | 7 (23.3%) | 2 (10.0%) | 9 (18.0%) |  |
| white | 23 (76.7%) | 18 (90.0%) | 41 (82.0%) |  |
| Education |  |  |  | 0.764 |
| high school or less | 5 (16.7%) | 4 (20.0%) | 9 (18.0%) |  |
| college or higher | 25 (83.3%) | 16 (80.0%) | 41 (82.0%) |  |
| Marital status |  |  |  | 0.386 |
| married/partnered | 19 (63.3%) | 15 (75.0%) | 34 (68.0%) |  |
| unmarried/unpartnered | 11 (36.7%) | 5 (25.0%) | 16 (32.0%) |  |
| Average duration of osteoarthritis (months) | 51.57 (55.44) | 80.20 (62.94) | 63.02 (59.63) | 0.043* |
| Kellgren-Lawrence score (index knee) |  |  |  | 0.868 |
| 0-1 | 4 (13.3%) | 3 (15.0%) | 7 (14.0%) |  |
| ≥ 2 | 26 (86.7%) | 17 (85.0%) | 43 (86.0%) |  |
| HPTh, knee (°C) | 41.38 (3.25) | 39.95 (3.27) | 40.81 (3.30) | 0.118 |
| HPTo, knee (°C) | 46.37 (1.95) | 45.21 (2.54) | 45.91 (2.26) | 0.087 |
| HPTh, arm (°C) | 40.07 (3.38) | 38.91 (3.41) | 39.60 (3.41) | 0.166 |
| HPTo, arm (°C) | 45.98 (2.29) | 44.62 (3.03) | 45.43 (2.67) | 0.115 |
| PPTh, medial knee (kgf) | 3.35 (1.47) | 2.20 (0.80) | 2.89 (1.36) | 0.004* |
| PPTh, trapezius (kgf) | 3.58 (1.95) | 2.37 (0.57) | 3.10 (1.65) | 0.009* |
| Punctate mechanical pain, patella (average series of 10 in patella punctate mechanical, score: 0-100) | 7.38 (10.32) | 9.18 (16.99) | 8.10 (13.26) | 0.944 |
| Temporal summation of pain, patella (series of 10 - single trial) | 2.82 (6.38) | 5.17 (16.01) | 3.76 (11.18) | 0.432 |
| Punctate mechanical pain, hand (average series of 10 in patella punctate mechanical, score: 0-100) | 5.82 (9.46) | 3.67 (5.70) | 4.96 (8.16) | 0.730 |
| Temporal summation of pain, hand (series of 10 - single trial) | 3.32 (7.26) | 2.40 (5.30) | 2.95 (6.50) | 0.890 |
| CPM (PPTh at 60 seconds - PPTh at pre-CPM) | 4.26 (2.27) | 2.95 (0.82) | 3.74 (1.93) | 0.019* |
| Cold pain intensity (score: 0-100) | 71.27 (26.07) | 77.25 (29.00) | 73.66 (27.15) | 0.266 |
| Depression (CES-D) | 17.53 (4.99) | 16.50 (5.53) | 17.12 (5.18) | 0.383 |
| Pain catastrophizing (PCS) | 8.77 (8.40) | 8.35 (6.77) | 8.60 (7.72) | 0.691 |
| COVID stress (CSS) | 10.13 (11.54) | 8.45 (10.28) | 9.46 (10.98) | 0.619 |
| *β coefficients (HbO) by fNIRS channel in response to punctate stimuli* |  |  |  |  |
| S01-D01 | 0.02 (3.72) | -0.29 (2.95) | -0.06 (3.48) | 0.977 |
| S01-D03 | -1.24 (2.62) | -0.20 (2.52) | -0.90 (2.60) | 0.492 |
| S02-D02 | 0.57 (2.65) | 1.11 (4.68) | 0.77 (3.46) | 0.655 |
| S02-D04 | 1.23 (3.15) | 1.23 (5.56) | 1.23 (4.18) | 0.210 |
| S03-D01 | 0.27 (2.46) | -0.53 (3.18) | -0.03 (2.71) | 0.743 |
| S03-D03 | 0.74 (3.89) | -0.43 (4.69) | 0.34 (4.14) | 0.646 |
| S03-D05 | 0.25 (3.56) | -0.70 (3.20) | -0.16 (3.39) | 0.572 |
| S04-D02 | -0.42 (4.25) | -1.02 (4.36) | -0.65 (4.24) | 0.684 |
| S04-D04 | -0.27 (3.05) | 0.39 (3.44) | -0.05 (3.12) | 1.000 |
| S04-D06 | 0.42 (2.28) | 0.04 (3.74) | 0.29 (2.79) | 0.758 |
| S05-D03 | 0.61 (2.79) | 4.10 (3.95) | 1.89 (3.60) | 0.176 |
| S05-D05 | -0.28 (4.79) | 1.49 (3.68) | 0.31 (4.45) | 0.854 |
| S05-D07 | 0.61 (3.25) | 1.46 (2.07) | 0.93 (2.85) | 0.161 |
| S06-D04 | 0.40 (3.16) | 2.13 (3.37) | 0.98 (3.28) | 0.127 |
| S06-D06 | -0.97 (4.11) | 1.42 (1.66) | -0.03 (3.53) | 0.003* |
| S06-D08 | -1.69 (5.90) | 1.50 (2.62) | -0.31 (4.97) | 0.015* |
| S07-D05 | 0.62 (5.47) | 0.16 (1.93) | 0.46 (4.47) | 0.910 |
| S07-D07 | -0.04 (4.62) | -0.44 (2.01) | -0.17 (3.93) | 0.704 |
| S08-D06 | -1.22 (4.86) | -1.46 (5.55) | -1.29 (4.97) | 0.912 |
| S08-D08 | -0.17 (3.23) | -1.52 (4.24) | -0.66 (3.62) | 0.454 |
| *β coefficients (HbO) by fNIRS ROIs in response to punctate stimuli* |  |  |  |  |
| Right dorsolateral prefrontal cortex (rdlPFC) | -0.47 (2.41) | -0.12 (2.88) | -0.34 (2.56) | 0.820 |
| Medial prefrontal cortex (mPFC) | 0.18 (2.27) | 0.63 (1.24) | 0.37 (1.90) | 0.675 |
| Left dorsolateral prefrontal cortex (ldlPFC) | 0.35 (3.49) | 0.49 (1.29) | 0.40 (2.90) | 0.297 |
| Right somatosensory cortex (rS1) | -0.38 (3.68) | 0.63 (2.64) | 0.02 (3.31) | 0.761 |
| Left somatosensory cortex (lS1) | -1.12 (3.32) | 0.77 (3.06) | -0.34 (3.31) | 0.017* |
| *β coefficients (HbO) by fNIRS channel in response to heat stimuli* |  |  |  |  |
| S01-D01 | 2.33 (6.31) | -0.33 (3.14) | 1.48 (5.57) | 0.200 |
| S01-D03 | 1.26 (3.15) | -1.01 (1.76) | 0.48 (2.93) | 0.016* |
| S02-D02 | 0.59 (4.08) | 0.80 (1.26) | 0.67 (3.27) | 0.270 |
| S02-D04 | 0.89 (5.15) | 0.18 (3.65) | 0.61 (4.57) | 0.971 |
| S03-D01 | 0.03 (3.39) | -1.06 (3.36) | -0.38 (3.35) | 0.571 |
| S03-D03 | 1.45 (4.37) | 0.68 (3.52) | 1.17 (4.02) | 0.482 |
| S03-D05 | -0.49 (2.98) | -1.34 (2.33) | -0.86 (2.71) | 0.630 |
| S04-D02 | -0.64 (4.41) | -1.30 (3.36) | -0.90 (3.99) | 0.622 |
| S04-D04 | -0.50 (2.36) | 1.99 (1.46) | 0.45 (2.38) | 0.006* |
| S04-D06 | 0.33 (3.40) | 1.46 (2.75) | 0.69 (3.20) | 0.446 |
| S05-D03 | 1.57 (4.21) | -0.49 (2.57) | 0.72 (3.68) | 0.495 |
| S05-D05 | 1.30 (5.74) | 0.24 (4.15) | 0.89 (5.10) | 0.885 |
| S05-D07 | -1.25 (2.63) | -1.42 (2.80) | -1.32 (2.64) | 0.907 |
| S06-D04 | 0.55 (5.45) | -1.64 (2.94) | -0.25 (4.76) | 0.116 |
| S06-D06 | -1.76 (6.07) | -1.63 (3.10) | -1.70 (4.83) | 0.355 |
| S06-D08 | -2.25 (8.68) | -1.14 (3.90) | -1.74 (6.84) | 0.903 |
| S07-D05 | -0.19 (3.27) | 1.75 (2.96) | 0.51 (3.24) | 0.126 |
| S07-D07 | 0.04 (3.75) | 2.34 (3.84) | 0.90 (3.89) | 0.129 |
| S08-D06 | 0.27 (3.30) | -0.67 (4.78) | -0.06 (3.81) | 0.225 |
| S08-D08 | 0.25 (3.77) | 0.05 (3.61) | 0.17 (3.65) | 0.330 |
| *β coefficients (HbO) by fNIRS ROIs in response to heat stimuli* |  |  |  |  |
| Right dorsolateral prefrontal cortex (rdlPFC) | 1.35 (2.58) | -1.21 (2.66) | 0.32 (2.87) | 0.012* |
| Medial prefrontal cortex (mPFC) | 0.90 (2.72) | -0.62 (2.86) | 0.25 (2.84) | 0.152 |
| Left dorsolateral prefrontal cortex (ldlPFC) | -0.58 (2.86) | 1.02 (2.12) | 0.04 (2.69) | 0.079 |
| Right somatosensory cortex (rS1) | 0.05 (2.67) | -0.73 (3.47) | -0.27 (3.01) | 0.958 |
| Left somatosensory cortex (lS1) | -0.44 (4.28) | -1.07 (3.60) | -0.71 (3.97) | 0.134 |

*Abbreviation.* CES-D, Center for Epidemiologic Studies Depression Scale; CPM, conditioned pain modulation; CSS, COVID stress scales; PCS, pain catastrophizing scale; PPTh, pressure pain threshold; HPTh, heat pain threshold, HPTo, heat pain tolerance; tDCS, transcranial direct current stimulation; MBM, mindfulness-based meditation; ROI, region of interest; HbO, oxygenated hemoglobin concentration

Significance levels: **p* < 0.05

### **Supplemental Table 4.** LCGA classification results for NRS: active tDCS + sham MBM

|  | *n* (%) or mean ± standard deviation | | |  |
| --- | --- | --- | --- | --- |
|  | Non-responder (*n* = 17) | Responder (*n* = 33) | Total (*n* = 50) | *p*-value |
| Age, years | 70.35 (6.03) | 66.88 (7.86) | 68.06 (7.42) | 0.112 |
| Body mass index (kg/m^2^) | 28.26 (7.29) | 29.43 (6.85) | 29.04 (6.95) | 0.287 |
| Gender |  |  |  | 0.318 |
| male | 7 (41.2%) | 9 (27.3%) | 16 (32.0%) |  |
| female | 10 (58.8%) | 24 (72.7%) | 34 (68.0%) |  |
| Race |  |  |  | 0.744 |
| non-white | 2 (11.8%) | 5 (15.2%) | 7 (14.0%) |  |
| white | 15 (88.2%) | 28 (84.8%) | 43 (86.0%) |  |
| Education |  |  |  | 0.296 |
| high school or less | 2 (11.8%) | 8 (24.2%) | 10 (20.0%) |  |
| college or higher | 15 (88.2%) | 25 (75.8%) | 40 (80.0%) |  |
| Marital status |  |  |  | 0.775 |
| married/partnered | 13 (76.5%) | 24 (72.7%) | 37 (74.0%) |  |
| unmarried/unpartnered | 4 (23.5%) | 9 (27.3%) | 13 (26.0%) |  |
| Average duration of osteoarthritis (months) | 37.71 (48.85) | 33.39 (35.30) | 34.860 (39.97) | 0.967 |
| Kellgren-Lawrence score (index knee) |  |  |  |  |
| 0-1 | 3 (17.6%) | 3 (9.1%) | 6 (12.0%) | 0.378 |
| ≥ 2 | 14 (82.4%) | 30 (90.9%) | 44 (88.0%) |  |
| HPTh, knee (°C) | 41.88 (4.27) | 39.11 (2.95) | 40.05 (3.66) | 0.022* |
| HPTo, knee (°C) | 46.63 (2.72) | 45.03 (2.22) | 45.57 (2.50) | 0.020* |
| HPTh, arm (°C) | 39.09 (2.99) | 37.47 (2.55) | 38.02 (2.79) | 0.054 |
| HPTo, arm (°C) | 45.23 (4.00) | 43.89 (3.14) | 44.34 (3.48) | 0.124 |
| PPTh, medial knee (kgf) | 3.02 (1.58) | 2.56 (1.38) | 2.71 (1.45) | 0.506 |
| PPTh, trapezius (kgf) | 2.73 (1.02) | 2.72 (1.55) | 2.73 (1.38) | 0.525 |
| Punctate mechanical pain, patella (average series of 10 in patella punctate mechanical, score: 0-100) | 6.94 (8.81) | 9.64 (13.27) | 8.72 (11.92) | 0.942 |
| Temporal summation of pain, patella (series of 10 - single trial) | 3.94 (8.69) | 3.30 (9.60) | 3.52 (9.21) | 0.560 |
| Punctate mechanical pain, hand (average series of 10 in patella punctate mechanical, score: 0-100) | 8.59 (13.14) | 8.59 (15.17) | 8.59 (14.38) | 0.268 |
| Temporal summation of pain, hand (series of 10 - single trial) | 5.32 (12.76) | 2.42 (5.86) | 3.41 (8.80) | 0.418 |
| CPM (PPTh at 60 seconds - PPTh at pre-CPM) | 2.93 (0.86) | 3.23 (1.49) | 3.13 (1.31) | 0.878 |
| Cold pain intensity (score: 0-100) | 72.65 (27.73) | 66.06 (30.69) | 68.30 (29.60) | 0.634 |
| Depression (CES-D) | 18.00 (6.59) | 19.36 (8.81) | 18.90 (8.08) | 0.532 |
| Pain catastrophizing (PCS) | 9.77 (7.46) | 13.27 (11.48) | 12.08 (10.35) | 0.412 |
| COVID stress (CSS) | 12.18 (10.89) | 10.03 (10.99) | 10.76 (10.89) | 0.365 |
| *β coefficients (HbO) by fNIRS channel in response to punctate stimuli* |  |  |  |  |
| S01-D01 | 0.78 (3.23) | 0.02 (4.17) | 0.40 (3.65) | 0.496 |
| S01-D03 | 1.05 (2.33) | -0.30 (2.52) | 0.27 (2.50) | 0.094 |
| S02-D02 | 0.59 (2.58) | -1.98 (5.45) | -0.95 (4.62) | 0.316 |
| S02-D04 | 0.36 (2.70) | -2.64 (6.54) | -1.42 (5.46) | 0.139 |
| S03-D01 | -2.16 (2.11) | -0.34 (3.08) | -1.30 (2.69) | 0.211 |
| S03-D03 | -0.19 (2.66) | -0.47 (3.51) | -0.32 (3.01) | 0.839 |
| S03-D05 | 0.48 (2.55) | -0.32 (3.70) | 0.04 (3.21) | 0.451 |
| S04-D02 | -0.64 (3.93) | 0.41 (4.80) | 0.03 (4.47) | 0.365 |
| S04-D04 | 0.42 (2.69) | -0.56 (1.97) | -0.10 (2.33) | 0.414 |
| S04-D06 | 0.29 (3.28) | -0.36 (2.45) | -0.11 (2.75) | 0.614 |
| S05-D03 | 0.23 (2.11) | -0.68 (6.20) | -0.17 (4.24) | 0.791 |
| S05-D05 | 0.88 (5.19) | -2.80 (7.10) | -0.88 (6.32) | 0.325 |
| S05-D07 | -1.89 (3.49) | -0.50 (3.15) | -1.20 (3.32) | 0.412 |
| S06-D04 | -3.11 (6.83) | -0.26 (4.63) | -1.47 (5.72) | 0.264 |
| S06-D06 | 0.84 (2.89) | 0.34 (4.90) | 0.55 (4.10) | 0.330 |
| S06-D08 | 0.43 (3.16) | 1.47 (4.89) | 1.09 (4.32) | 0.506 |
| S07-D05 | 0.28 (1.76) | 2.28 (5.31) | 1.38 (4.14) | 0.569 |
| S07-D07 | 0.49 (3.11) | 0.21 (3.22) | 0.33 (3.12) | 0.781 |
| S08-D06 | 1.00 (3.25) | -0.65 (4.30) | 0.14 (3.83) | 0.673 |
| S08-D08 | 0.59 (2.59) | -0.72 (4.80) | -0.17 (4.02) | 0.230 |
| *β coefficients (HbO) by fNIRS ROIs in response to punctate stimuli* |  |  |  |  |
| right dorsolateral prefrontal cortex (rdlPFC) | 0.18 (2.52) | -0.41 (2.83) | -0.15 (2.67) | 0.245 |
| medial prefrontal cortex (mPFC) | 0.07 (2.14) | -1.30 (3.29) | -0.72 (2.90) | 0.271 |
| left dorsolateral prefrontal cortex (ldlPFC) | -0.38 (1.41) | 0.60 (2.77) | 0.21 (2.34) | 0.150 |
| right somatosensory cortex (rS1) | -0.07 (2.86) | -0.92 (2.73) | -0.60 (2.77) | 0.442 |
| left somatosensory cortex (lS1) | 0.32 (2.00) | 0.27 (2.95) | 0.29 (2.59) | 0.688 |
| *β coefficients (HbO) by fNIRS channel in response to heat stimuli* |  |  |  |  |
| S01-D01 | -0.86 (2.94) | -0.42 (3.66) | -0.65 (3.24) | 0.538 |
| S01-D03 | 0.24 (2.56) | -0.49 (2.29) | -0.16 (2.41) | 0.974 |
| S02-D02 | 1.62 (4.00) | 2.06 (3.09) | 1.89 (3.37) | 0.469 |
| S02-D04 | 1.11 (3.96) | 2.07 (2.77) | 1.66 (3.30) | 0.265 |
| S03-D01 | -0.60 (3.65) | 1.15 (1.70) | 0.22 (2.96) | 0.211 |
| S03-D03 | -0.37 (3.43) | 0.26 (3.10) | -0.06 (3.22) | 0.778 |
| S03-D05 | -0.28 (2.30) | 0.06 (2.89) | -0.09 (2.62) | 0.754 |
| S04-D02 | -1.05 (4.18) | -0.71 (3.47) | -0.85 (3.72) | 0.951 |
| S04-D04 | 1.42 (3.75) | 0.90 (3.10) | 1.15 (3.34) | 0.806 |
| S04-D06 | 1.44 (3.04) | 0.88 (2.19) | 1.13 (2.55) | 0.511 |
| S05-D03 | 1.09 (3.03) | 0.28 (0.77) | 0.76 (2.36) | 0.845 |
| S05-D05 | 0.80 (3.03) | -1.69 (7.75) | -0.39 (5.79) | 0.580 |
| S05-D07 | -2.29 (7.12) | -1.42 (5.04) | -1.85 (6.04) | 0.818 |
| S06-D04 | -4.15 (9.18) | -3.37 (7.79) | -3.72 (8.30) | 0.726 |
| S06-D06 | 0.39 (2.33) | 0.46 (2.00) | 0.43 (2.10) | 0.966 |
| S06-D08 | -0.31 (2.93) | -0.02 (3.25) | -0.13 (3.09) | 0.964 |
| S07-D05 | 0.12 (1.24) | 1.25 (2.69) | 0.81 (2.27) | 0.378 |
| S07-D07 | 0.15 (2.07) | 1.33 (3.02) | 0.82 (2.67) | 0.403 |
| S08-D06 | 0.94 (3.40) | -0.75 (2.28) | 0.06 (2.92) | 0.291 |
| S08-D08 | 1.38 (3.45) | -0.95 (3.02) | 0.06 (3.37) | 0.047* |
| *β coefficients (HbO) by fNIRS ROIs in response to heat stimuli* |  |  |  |  |
| right dorsolateral prefrontal cortex (rdlPFC) | -0.45 (2.02) | -0.01 (1.86) | -0.21 (1.92) | 0.354 |
| medial prefrontal cortex (mPFC) | 0.14 (1.71) | -0.82 (5.47) | -0.43 (4.34) | 0.643 |
| left dorsolateral prefrontal cortex (ldlPFC) | -0.89 (3.38) | 0.95 (3.04) | 0.22 (3.26) | 0.352 |
| right somatosensory cortex (rS1) | -0.07 (3.58) | 0.81 (2.34) | 0.47 (2.87) | 0.751 |
| left somatosensory cortex (lS1) | 0.41 (2.65) | -0.23 (1.78) | 0.02 (2.16) | 0.473 |

*Abbreviation.* CES-D, Center for Epidemiologic Studies Depression Scale; CPM, conditioned pain modulation; CSS, COVID stress scales; PCS, pain catastrophizing scale; PPTh, pressure pain threshold; HPTh, heat pain threshold, HPTo, heat pain tolerance; tDCS, transcranial direct current stimulation; MBM, mindfulness-based meditation; ROI, region of interest; HbO, oxygenated hemoglobin concentration

Significance levels: **p* < 0.05
